# Spatiotemporal Trends in Group A Streptococcal Pharyngitis in the United States

**DOI:** 10.1101/2023.11.16.23298647

**Authors:** Madeleine C. Kline, Stephen M. Kissler, Lilith K. Whittles, Michael L. Barnett, Yonatan H. Grad

**Author notes:** Corresponding author: Yonatan H. Grad, Harvard T.H. Chan School of Public Health, 665 Huntington Ave, Boston, MA 02115.

## Abstract

**Background:** Group A *Streptococcus* (GAS) causes an estimated 5.2 million outpatient visits for pharyngitis annually in the United States (U.S.) with incidence peaking in winter, but the annual spatiotemporal pattern of GAS pharyngitis across the U.S. is poorly characterized.

**Methods:** We used outpatient claims data from individuals with private medical insurance between 2010-2018 to quantify GAS pharyngitis visit rates across U.S. census regions, subregions, and states. We evaluated seasonal and age-based patterns of geographic spread and the association between school start dates and the summertime upward inflection in GAS visits.

**Results:** The South had the most visits per person (yearly average 39.11 visits per 1000 people, 95% CI: 36.21-42.01), and the West had the fewest (yearly average 17.63 visits per 1000 people, 95% CI: 16.76-18.49). Visits increased earliest in the South and in school-age children. Differences in visits between the South and other regions were most pronounced in the late summer through early winter. Visits peaked earliest in central southern states, in December to January, and latest on the coasts, in March. The onset of the rise in GAS pharyngitis visits correlated with, but preceded, average school start times.

**Conclusions:** The burden and timing of GAS pharyngitis varied across the continental U.S., with the South experiencing the highest overall rates and earliest onset and peak in outpatient visits. Understanding the drivers of these regional differences in GAS pharyngitis will help in identifying and targeting prevention measures.

**Summary:** The incidence of streptococcal pharyngitis varies across the U.S., with highest rates in the South and lowest in the West. On average, the yearly summer increase in cases begins in the South and progresses outwards to adjacent states.

## INTRODUCTION

Group A *Streptococcus* (GAS; *Streptococcus pyogenes*) is a common human bacterial pathogen that causes a diverse spectrum of disease. GAS pharyngitis, known commonly as “strep throat,” is responsible for an estimated 5.2 million outpatient visits and 2.8 million antibiotic prescriptions each year in the U.S., accounting for 5.9% of all outpatient antibiotic prescriptions in children ages 3-9 years.^1^ According to the Centers for Disease Control and Prevention (CDC), GAS accounts for 20-30% of pharyngitis cases in children and 5-15% of pharyngitis cases in adults.^2^ GAS pharyngitis is more common in the winter and spring months,^2–4^ but its seasonality and geography in the U.S. remains poorly characterized.

Other common respiratory conditions have well-characterized spatiotemporal trends: epidemic waves of influenza usually start in the southern U.S., and seasonal peaks of respiratory syncytial virus (RSV) are typically earliest in Florida.^5,6^ Environmental factors such as mean absolute humidity, vapor pressure, minimum temperature, and precipitation have been proposed as transmission modifiers for these viruses.^5,6^ School-aged children play an important role in community influenza transmission and in the transmission and introduction of RSV into households.^7,8^ Whether these factors similarly impact GAS pharyngitis transmission remains unclear.

To address the need for improved characterization of GAS epidemiology in the U.S., we used outpatient insurance claims data from private insurers to assess geographic patterns of visits for GAS pharyngitis over the course of the year and to evaluate the association with the start of the school year.

## METHODS

### Study Population and Data Source

Retrospective outpatient claims data were extracted from the Merative™ MarketScan® Commercial Database, which is a convenience sample of 16.6-36.4 million annually-enrolled privately-insured individuals (5.1-11.6% of the total U.S. population, depending on the year).^9^ These data include claims from outpatient care, pharmacy, and enrollment data from large health plans and employers covering employees and their families. The sample was restricted to individuals who were continuously enrolled for an entire year between 2010-2018 and included information on age group, sex, and state (**Table 1, Figure S1, Figure S2**).

**Table 1:** Study Population Characteristics. This table reports the mean and range in the annual number of individuals included in the MarketScan database (2010-2018) by sex, age group, region, and subregion.

| Category | Average Membership x10<sup>6</sup> (Range) | % |
| --- | --- | --- |
| **Total** | 26.7 (16.6-36.42) | 100 |
| Sex |  |  |
| Male | 12.95 (8.07-17.77) | 48.49 |
| Female | 13.75 (8.53-18.66) | 51.51 |
| Age Group |  |  |
| 0-4 | 1.5 (0.92-2.08) | 5.60 |
| 5-9 | 1.79 (1.07-2.49) | 6.71 |
| 10-19 | 4.15 (2.46-5.78) | 15.55 |
| 20-29 | 3.53 (2.41-4.8) | 13.22 |
| 30-39 | 3.9 (2.48-5.26) | 14.62 |
| 40-49 | 4.74 (2.81-6.58) | 17.77 |
| 50-59 | 5.21 (3.16-7.08) | 19.51 |
| 60-69 | 1.88 (1.29-2.4) | 7.03 |
| Region |  |  |
| Midwest | 6.18 (4.01-8.95) | 23.14 |
| Northeast | 4.92 (2.65-7.09) | 18.44 |
| South | 10.34 (6.78-13.14) | 38.75 |
| West | 5.25 (2.79-7.69) | 19.68 |
| Subregion |  |  |
| East North Central | 4.86 (3-7.3) | 18.21 |
| East South Central | 1.7 (1.04-2.1) | 6.37 |
| Middle Atlantic | 3.76 (1.96-5.69) | 14.08 |
| Mountain West | 1.6 (0.94-2.24) | 6.00 |
| New England | 1.16 (0.65-1.72) | 4.36 |
| Pacific West | 3.65 (1.85-5.45) | 13.68 |
| South Atlantic | 5.3 (3.71-6.44) | 19.85 |
| West North Central | 1.32 (1-1.73) | 4.93 |
| West South Central | 3.34 (2.03-4.83) | 12.53 |

### Disease Incidence

Visits prompted by GAS pharyngitis were identified by mapping codes from the *International Classification of Diseases, Clinical Modification* ninth (ICD9) or tenth (ICD10) revision to Clinical Classification Software (CCS) codes (**Table S1**).^10^ Guidelines recommend GAS pharyngitis diagnosis by a rapid antigen detection test (RADT) or throat culture, rather than by clinical features alone.^2,11^ Visits were thus included if a diagnosis code indicating GAS pharyngitis was the first or second diagnosis billed for the visit.

Yearly, quarterly, and monthly visits per 1,000 people within the 48 continental U.S. states plus Washington D.C., 9 subregions, and 4 regions (according to the Census Regions and Divisions of the United States, **Tables S2 and S3**)^12^ were calculated by dividing the number of visits in the respective time period by the number of people enrolled during that period in the respective geographic division and multiplying by 1,000. There was no clear secular trend in visits across years (**Figure S3**), and thus visits were averaged across all 9 years of observation and 95% confidence intervals were calculated under the assumption of normally distributed errors. There were no clear differences in age distributions by region; consequently, visits from all ages were included unless otherwise specified (**Figure S2**).

To account for potential differences between the age and sex distribution of the MarketScan data and the general population, visit counts were weighted by their census-determined proportion of the population. State-level population counts were obtained from the 2011-2015 American Community Survey (ACS) and accessed using the tidycensus R package^13^.

### Statistical Analyses

#### Regional Significance Testing

To assess regional differences in visit rates, yearly visits per person in each region were compared using Welch’s two sample t-test. To assess differences in the seasonality of GAS visits across regions, visits in each region and month were compared to all other regions in that month using Welch’s two sample t-test. Statistical significance was determined based on a significance level of 0.05 corrected for multiple hypothesis testing using the Bonferroni correction.

#### Seasonal Modeling

To characterize the seasonality in GAS pharyngitis visits by state or region, we fit sinusoids to data from the 9 years of observation using nonlinear least squares regression. Trends were modelled using the following equation:

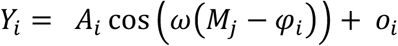

where *Y*_*i*_ is the number of visits per thousand people in state or region *i, A*_*i*_ is the amplitude for state/region *i* (difference between the maximum and minimum monthly visits per 1000 people in a year), 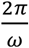 is the period (the time required for one full cycle of visits), *M*_*j*_ is the month of observation *j* (taking values from 1-12 with 1 corresponding to January 1^st^), *φ*_*i*_ is the phase (the horizontal shift in months, so that *φ*_*i*_ is the month in which the maximum visits per 1000 people occurs), and *o*_*i*_ is the baseline visit rate (i.e., the mean number of visits per 1000 people in location *i*). The period was fixed at 12 months 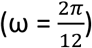. Estimates of the amplitude, phase, and offset were optimized using the *nls* function in R. The initial amplitude was specified as half the distance from the average maximum to minimum number of visits in the entire dataset. The initial phase was specified as the average month during which the maximum number of visits occurred. The initial offset was specified as the mean number of visits throughout the year. Confidence regions for sinusoid predictions were calculated via bootstrapping (see supplemental text for details).

#### Geographic Patterns

Population-weighted state centroids from 2010 (calculated by the U.S. Census Bureau) were extracted from the USpopcenters R package.^14,15^ We calculated subregion centroids using a population-weighted average of the state centroid. For the reference point, we used the population-weighted average between the East South Central and West South Central subregions, since their sinusoid phases had the earliest peaks of all the subregions and were very similar. We calculated the Euclidean distance from the reference point to the centroid of each state or subregion. We correlated the vector of distances for each state or subregion to the vector of sinusoid phases, representing peak timing, for each state or subregion using Pearson’s correlation coefficient.

#### School Start Time Correlation

Data on 2019 school start dates was obtained from the Pew Research Center.^16^ This dataset contains a 509-district sample of the >13,000 public school districts in the U.S., including the 10 largest school districts in states with at least 10, and the 100 largest school districts in the country, representing an estimated 36% of public-school students. We filtered the data to include only continental U.S. states, yielding 497 school districts. School districts were grouped at the state and subregion level. We calculated the dates at which GAS pharyngitis visits were at their nadir by taking the average of the month with the fewest visits in each state or subregion for each of the 9 years of observation. We calculated minimum visit dates by subregion in all age groups together, and separately in those aged 0-4, 5-19, under 19, and over 19. We determined the relationship between the average school start date and minimum visit date in each state or subregion via Pearson’s correlation coefficient. We repeated this analysis separately for those under and over 19 years of age. The 95% confidence interval on the correlation coefficient was calculated via bootstrapping (see supplemental text for further details).

## RESULTS

### Difference in Disease Burden

#### Yearly and Quarterly Differences

Among the four census regions, the South had the highest mean number of GAS pharyngitis visits per 1000 people per year (39.11, 95% CI: 36.21-42.01) and the West had the lowest (17.63, 95% CI: 16.76-18.49); (**Figure S3**). At a subregional level, the East South Central region had the highest mean number of GAS pharyngitis visits per 1000 people per year (48.38, 95% CI: 42.40-53.37), while the Pacific West had the lowest (12.39, 95% CI: 11.57-13.22); (**Figure S4, Table S4**). Mean GAS pharyngitis visit rates in the South and the West each differed significantly from those in all other regions (p < 0.05) (**Figure S3**).

Differences in age distributions across regions were insufficient to explain regional differences in GAS pharyngitis visits. The overall age distribution appears similar across all regions (**Figure S2, Table S5**). In the South, a relatively greater share of GAS pharyngitis visits was attributable to 0–4-year-olds than in other regions (26% in the South compared to 18% in the West), but this age group made up a similar proportion of the population in each region (5% in the South, 6% in the West). In all regions, individuals under 19 years old contributed the most GAS pharyngitis visits (range: 67% in the West – 77% in the Northeast). This group made up a similar fraction of each region’s overall population (range: 26.8% in the Northeast – 28.7% in the Midwest).

During each quarter of the year, the West maintained a lower mean number of visits per 1,000 people than the other three regions (**Table S6**). The differences in mean visits per 1,000 people between the South, the Northeast, and the Midwest were more pronounced in the summer/fall (July through December) than in the winter/spring (January through June) (**Figure S5, Table S5**).

#### Monthly Differences

To better resolve the seasonal patterns in GAS pharyngitis incidence, we compared monthly GAS pharyngitis visits across regions and subregions. Across regions, GAS pharyngitis visits were more common in the winter months, with visits nadiring in the summer months before inflecting upwards in early autumn and peaking in the first few months of the calendar year (**Figure 1**). In the South, for example, the January mean was 3.78 (95% CI: 3.36-4.21) visits per 1000 people, while in July the mean was 1.80 (95% CI: 1.67-1.93) visits per 1000 people. In the West, visits were much lower; the January mean was 1.76 (95% CI: 1.62-1.90) visits per 1000 people, while in July it was 0.98 (95% CI: 0.93-1.03). At a subregional level, the East South Central region had on average 4.70 (95% CI: 4.13-5.26) visits per 1000 people in January and 1.88 (95% CI: 1.71-2.06) visits per 1000 people in July, while the Pacific West had on average 1.2 (95% CI: 1.09-1.31) visits per 1000 people in January and 0.76 (95% CI: 0.71-0.81) visits per 1000 people in July (**Figure S6**). Comparisons across regions and subregions revealed statistically significant differences in monthly GAS pharyngitis incidence in the second half of the year (**Figure S7**). These trends in visits between subregions over the course of the year were reflected across all age groups. Throughout the year, most visits came from those under 19 years of age (**Figure S8**).

**Figure 1:**
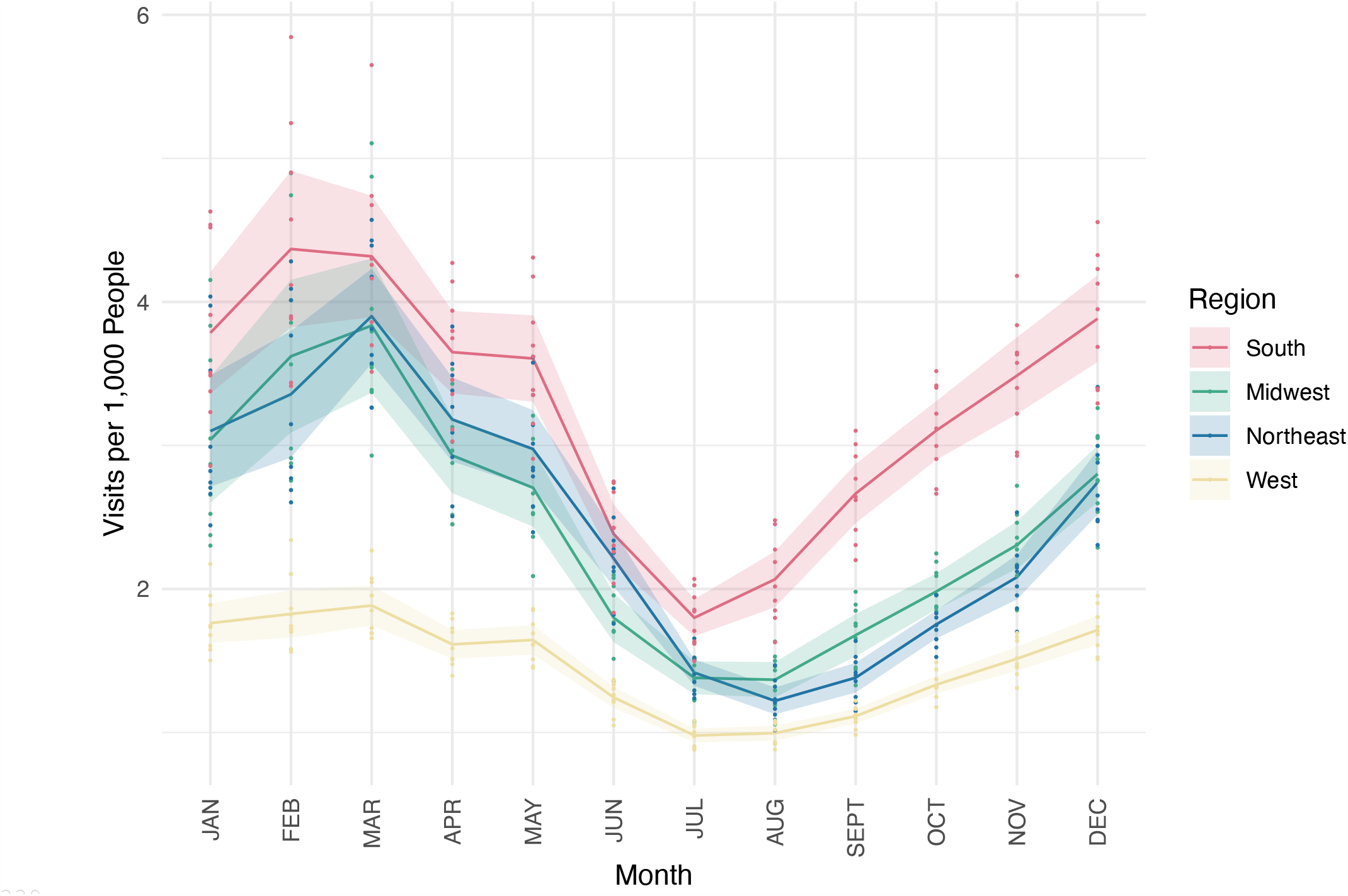
Average monthly GAS pharyngitis visits per 1,000 people by census region. Points depict the mean number of GAS pharyngitis visits observed in a given region (color), month (horizontal position), and year. The monthly mean values for each region are connected by solid lines. Shaded regions represent the 95% confidence intervals calculated assuming normally distributed errors pertaining to the monthly mean values, and thus depict year-to-year variation in GAS pharyngitis visit rates.

### Differences in Peak Timing

We estimated the timing of peak GAS pharyngitis incidence using sinusoidal models fit to monthly GAS pharyngitis visits at the state, subregional, and regional levels. The annual peak in GAS pharyngitis visits occurred earliest in the South, followed by peaks in adjacent states and through the Mountain West, with the latest peaks in coastal states (**Figure 2, Figure S9**). The states with the earliest peaks were in the East South Central (phase: 1.50, 95% CI 1.22-1.79, where 1.0 represents January 1^st^) and West South Central (phase: 1.49, 95% CI 1.20-1.77) subregions (**Figures S10 and S11**). The Mountain West had the next peak, (phase: 1.68, 95% CI 1.49-1.88) with the latest peaks on the coasts in March (Pacific West phase: 2.35, 95% CI 2.10-2.61; New England phase: 2.62, 95% CI 2.44-2.81). Louisiana (phase: 0.81, 95% CI 0.41-1.21) and Mississippi (phase: 0.86, 95% CI 0.43-1.3) peaked particularly early, in December (**Table S7**).

**Figure 2:**
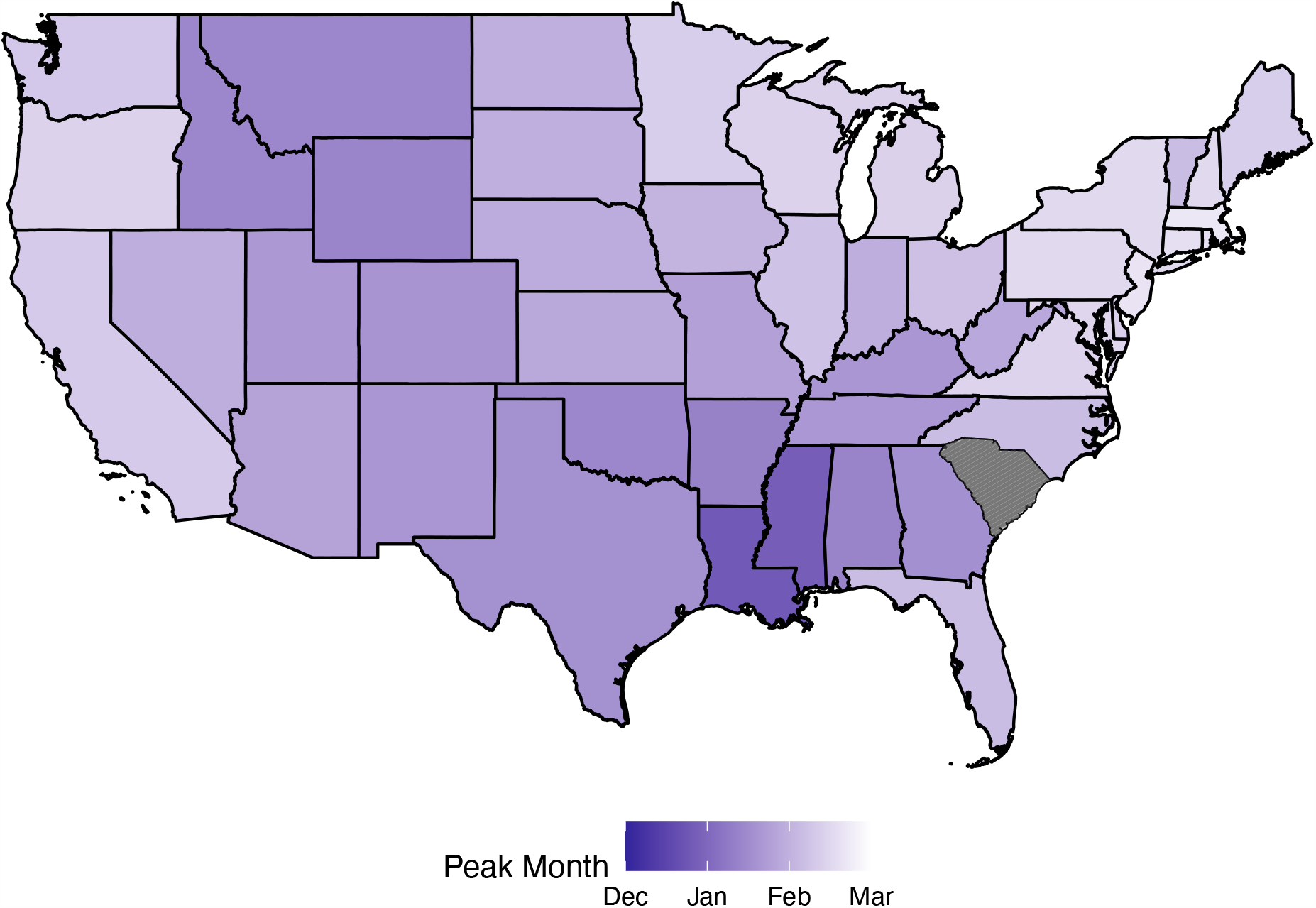
Timing of peak GAS pharyngitis visits by U.S. state. States are colored according to the estimated timing of peak GAS pharyngitis visits (phase of the sinusoidal fit to monthly GAS pharyngitis visits per 1,000 people across all ages). South Carolina is excluded from map according to the data use agreement. Darker colors indicate earlier peaks, and lighter colors indicate later peaks. See also the Supplemental Animation.

The annual peak time of GAS pharyngitis incidence correlated with geographic distance from the population-weighted centroid of the South (near Homer, Louisiana; latitude 32.83, longitude -93.01) (**Figures S12 and S13**). Greater distances from the southern reference point correlated with later peak times at both the subregional (Pearson’s r = 0.73, **Figure S12**) and state (Pearson’s r = 0.58, **Figure S13**) levels.

### School Start Date Analysis

School start dates correlated with the nadir in GAS pharyngitis visits, at state and subregional levels (state: Pearson’s *r* = 0.84, 95% CI: 0.82-0.86, subregional: Pearson’s *r* = 0.85, 95% CI: 0.76-0.92) (**Figure 3, Figures S14-16**). These correlations held in both the under 19 population (Pearson’s *r* = 0.82, 95% CI: 0.72-0.88) and the over 19 population (Pearson’s *r* = 0.89, 95% CI: 0.76-0.97). In all subregions and states, the nadir date preceded the school start date (mean number of days preceding by subregion: 36.22, range= 28-47 days, **Figure S14**; mean number of days preceding by state: 33.47, range = 14.9-48.5; **Figures S15 and S16**). The subregional minimum visit dates occurred first in those 5-19 years old, followed by those 0-4 years old, and then those over 19 years old (**Figure S17**).

**Figure 3:**
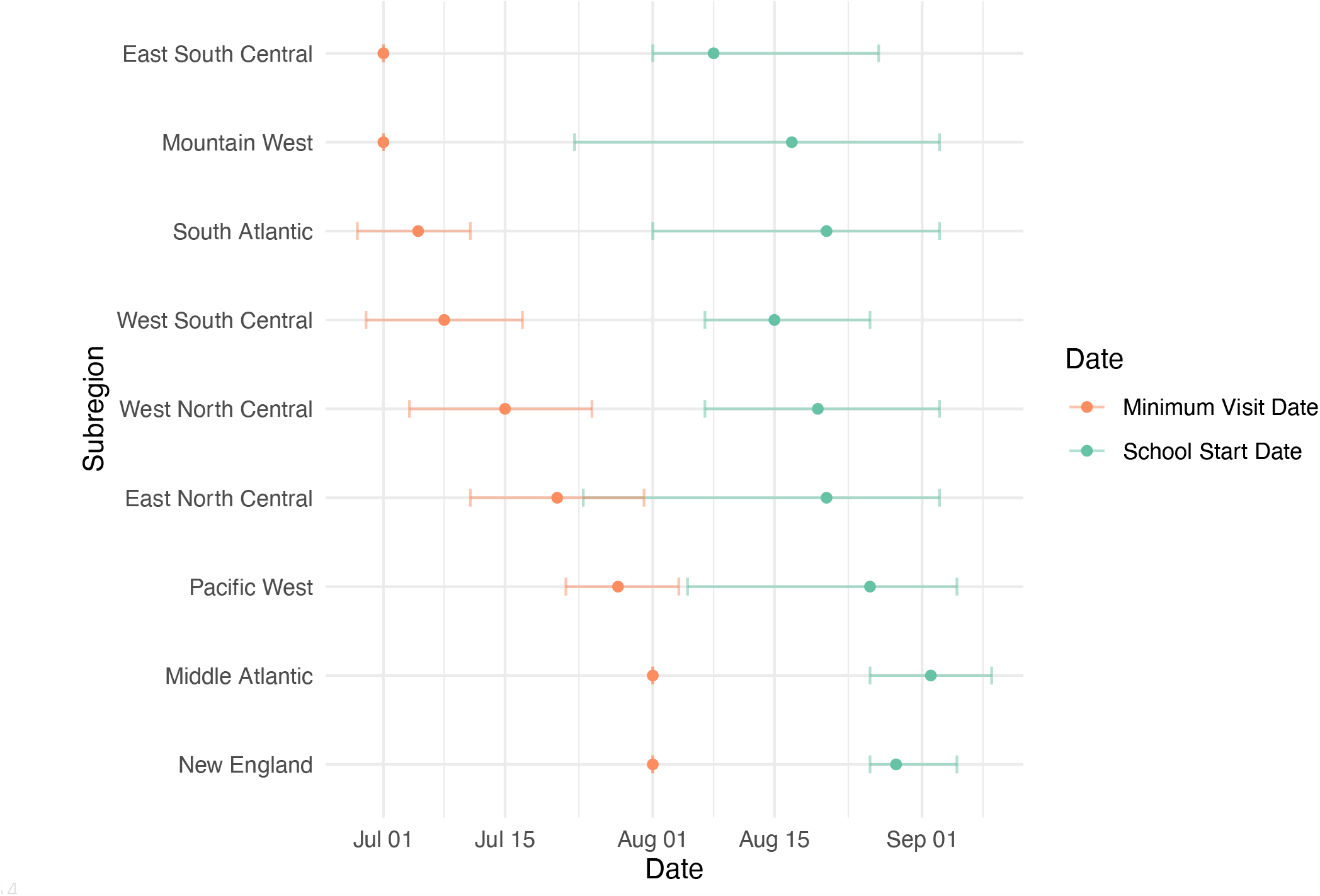
Relationship between subregion school start date and the nadir date of GAS pharyngitis visits. Estimated GAS pharyngitis nadir dates (orange points) with 95% confidence intervals assuming normally distributed errors (whiskers) are depicted alongside mean school start dates (green points) and ranges (whiskers) in that subregion.

## DISCUSSION

Our results expand on estimates of the seasonality and incidence of GAS pharyngitis in the U.S.^1,3,17^ Visits for GAS pharyngitis were higher in the South, and particularly the East South Central and West South Central subregions, than other regions throughout the year, especially from July to December. The Pacific West subregion documented the fewest GAS pharyngitis visits throughout the year. Annually, GAS pharyngitis incidence began to rise earliest in the same southern subregions that had the highest burden of disease, and peaked latest in coastal regions. The annual increase in visits was first observed in southern states, particularly in Louisiana and Mississippi, followed by the rest of the country roughly in order of increasing distance from the South. Visits increased first in the 5–19-year-old age group, followed by the 0–4-year-old age group and then the over 19-year-old age group across all subregions. This geographic pattern of increasing GAS pharyngitis cases correlated with, but preceded, school start dates.

The factors driving higher rates of GAS pharyngitis visits and the earliest rise in cases in the South are unclear. This could represent conditions that promote GAS pharyngitis, including those relating to poverty and social inequality^18^ or to other infections. GAS pharyngitis spatiotemporal patterns were similar to trends in influenza and RSV.^5^ Some of the drivers for those pathogens, such as climatic factors, may also contribute to the trends in GAS pharyngitis. Alternatively, and more speculatively, the extent of asymptomatic carriage of GAS (nearly 50% in children ages 3-12^4^) may result in false positive RADTs in the context of viral pharyngitis in hosts carrying GAS, such that the trends of viral infections may contribute to the observed trends of GAS pharyngitis.

Similarly, it is unclear what explains the low rates of GAS pharyngitis visits in the Pacific West subregion. This could be due to local differences in GAS strains and their propensity to cause pharyngitis, fewer susceptible hosts due to increased immunity from infection or a cross-protective pathogen, or environmental, social, or behavioral factors that reduce transmissibility. Systematic underreporting in private insurance claims in this region could also contribute to low observed rates.

Nadirs in GAS pharyngitis visits preceded and correlated with school start dates. Subregions in the South with earlier upticks in GAS pharyngitis visits also started school earlier than coastal subregions. Given that uptick dates preceded school start dates by over a month, it is unlikely that school attendance initiated transmission in a given season, but changes in contact patterns among children associated with the start of school may facilitate both the spread of GAS pharyngitis, like with other respiratory infections,^19^ and its ascertainment. The age pattern of epidemic onset, which occurred first in children aged 5-19 years, followed by those 0-4 years old and then adults over 19 years old, could implicate school-age children as an important infection-control target for GAS. Further examination of the relationship between rates of GAS pharyngitis and school closures enacted as part of COVID-19 response could help elucidate the role of school as a nidus of community spread of GAS pharyngitis.

Trends in GAS pharyngitis may indicate broader trends in GAS disease and differences in transmission mechanisms between different GAS clinical syndromes. Invasive GAS (iGAS) infections are highly morbid infections that occur when GAS invades a typically sterile body site, but their connection to GAS pharyngitis remains unclear.^20^ GAS necrotizing fasciitis has been shown not to follow seasonal patterns, although this may be due to small sample size, while overall iGAS cases are seasonal in the U.S.^3,21^ After reported iGAS cases in the U.S. declined by about 25% during the COVID-19 pandemic, there was an increase in iGAS infections in the U.S. in the 2022-2023 season.^22^ Whether this represents a particularly invasive strain, or increased population susceptibility resulting in higher rates of severe disease remains to be determined. Similarly, GAS pharyngitis rates also increased in the 2022-2023 season.^23^

Limitations to this study include that the data capturing GAS-related visits are a convenience sample of privately insured individuals in the U.S. Potential bias could also arise from geographic heterogeneity in insurance and care access and from restricting the dataset to individuals continuously enrolled over the course of the year, thus excluding people who changed insurers or moved states frequently. All visits where GAS pharyngitis was the primary or secondary diagnostic code were included, but there may have been differences in how providers across different states or hospital systems bill for this condition. School start data was not sampled comprehensively. Lastly, analyses at the state, subregion, and regional levels may not reflect local variation.

In conclusion, GAS pharyngitis was observed more and peaked earliest in the South compared to other regions. The Pacific West had fewer GAS-related visits than other subregions, and visits in coastal subregions peaked latest. Across all regions, visits increased first in those 5-19 years old, suggesting this age group is important for community transmission. As GAS pharyngitis causes a high burden of disease annually and is a major driver of antibiotic prescriptions in the U.S., the spatiotemporal patterns reported here can aid in designing effective surveillance programs and in allocating resources to reduce its incidence.

## Author contributions

M.C.K contributed to study design, conceptualization, and analysis, and drafted the manuscript. S.M.K contributed to study design, conceptualization, analysis and edited the manuscript. L.K.W. and M.L.B contributed to methodology and edited the manuscript. Y.H.G contributed to study design and conceptualization, edited the manuscript, and supervised the work.

## Funding sources

M.C.K. was supported by [grant award number T32GM144273] from the National Institute of General Medical Sciences. S.M.K. received funding from NIH T32 training grant 2 T32 AI 7535-21 A1. The content is solely the responsibility of the authors and does not necessarily represent the official views of the National Institute of General Medical Sciences or the National Institutes of Health. L.K.W. acknowledges funding from the Wellcome Trust [grant number 218669/Z/19/Z] and from the MRC Centre for Global Infectious Disease Analysis (reference MR/X020258/1), funded by the UK Medical Research Council (MRC). This UK funded award is carried out in the frame of the Global Health EDCTP3 Joint Undertaking. This project has been funded (in part) by contract 200-2016-91779 with the Centers for Disease Control and Prevention. Disclaimer: The findings, conclusions, and views expressed are those of the author(s) and do not necessarily represent the official position of the Centers for Disease Control and Prevention (CDC).

## Data availability

We cannot share disaggregated MarketScan or school start date data. MarketScan data are available via purchase of a commercial license. School start data was obtained via request to Pew Research Center. All code is available at https://github.com/mkline1/StrepPharyngitis_Public

## Conflicts of interest

No reported conflicts.

## Notes

### Competing Interest Statement

The authors have declared no competing interest.

