## Supplemental text, tables, and figures for "Spatiotemporal Trends in Group A Streptococcal Pharyngitis in the United States"

**SUPPLEMENTAL MATERIALS**
All code available at https://github.com/mkline1/StrepPharyngitis_Public

**Supplementary Text:**

*Sinusoid model details*

The phase $\varphi$ represents the month during which the peak in visits occurred, which is 6 months before and after the month with the minimum number of visits according to sinusoid structure with a 12-month period. We mapped months to integers by taking the phase modulo 12. Taking each integer value to be the first day of the specified month (e.g., 1 is January 1$^{st}$), the phases fell between 0-11.99 where 0 is the first day of December and 11.99 is the last day of November.

Confidence regions for the sinusoidal fits were calculated via bootstrapping to obtain a conservative confidence region estimate: 1000 samples were drawn from normal distributions centered around each of the 3 fitted sinusoid parameters (amplitude, phase, and offset) and with standard deviation equal to the standard error estimated by the model for each parameter. Sinusoids were generated using each of the 1000 sets of 3 bootstrapped parameters. Bounds for the 95% confidence regions of the mean visits per 1000 people in each month were then extracted as the 2.5$^{th}$ and 97.5$^{th}$ quantiles across this set of 1000 generated sinusoids.

*Pearson correlation confidence interval*

The confidence interval of the correlation coefficient was calculated via bootstrapping; 9 samples were randomly drawn with replacement from each subregion and averaged to get a bootstrapped mean minimum visit date in each subregion or state 1000 times. Similarly, the number of samples corresponding to the number of districts in a given state or subregion were drawn from the dataset of school starts with replacement and averaged to get a bootstrap mean school start date in each subregion or state 1000 times. These 1000 bootstrapped mean vectors were then correlated with one another using Pearson's correlation coefficient, and the 2.5$^{th}$, 50$^{th}$, and 97.5$^{th}$ percentiles were taken to get the average correlation and 95% confidence interval.

**Supplementary Table 1: CCS to ICD Code Mapping**

| ICD REVISION | CCS CATEGORY | CCS CATEGORY DESCRIPTION | ICD CODE | ICD CODE DESCRIPTION |
|---|---|---|---|---|
| ICD-9 | 126 | Ot up rsp in | 340 | STREP SORE THROAT |
| ICD-10 | RSP006 | Other specified upper respiratory infections | J020 | Streptococcal pharyngitis |

**Supplementary Table 2: Regions with constituent states**

| Region | States |
|---|---|
| Northeast | Connecticut, Massachusetts, Maine, New Hampshire, Rhode Island, Vermont, New Jersey, New York, Pennsylvania |
| Midwest | Illinois, Indiana, Michigan, Ohio, Wisconsin, Iowa, Kansas, Minnesota, Missouri, Nebraska, North Dakota, South Dakota |
| South | Delaware, Florida, Georgia, Maryland, North Carolina, South Carolina, Virginia, West Virginia, Alabama, Kentucky, Mississippi, Tennessee, Arkansas, Louisiana, Oklahoma, Texas |
| West | Arizona, Colorado, Idaho, Montana, Nevada, New Mexico, Utah, Wyoming, Alaska, California, Hawaii, Oregon, Washington |

**Supplementary Table 3: Subregions with constituent states**

| Subregion | States |
|---|---|
| New England | Connecticut, Maine, Massachusetts, New Hampshire, Rhode Island, Vermont |
| Middle Atlantic | New Jersey, New York, Pennsylvania |
| East North Central | Indiana, Illinois, Michigan, Ohio, Wisconsin |
| West North Central | Iowa, Kansas, Minnesota, Missouri, Nebraska, North Dakota, South Dakota |
| South Atlantic | Delaware, Washington DC, Florida, Georgia, Maryland, North Carolina, South Carolina, Virginia, West Virginia |
| East South Central | Alabama, Kentucky, Mississippi, Tennessee |
| West South Central | Arkansas, Louisiana, Oklahoma, Texas |
| Mountain West | Arizona, Colorado, Idaho, New Mexico, Montana, Utah, Nevada, Wyoming |
| Pacific West | California, Oregon, Washington |

**Supplementary Table 4:** Statistical comparisons in visits per 1000 people per year by subregion

| Subregion Comparison | p-value | Difference in visits per 1000 people per year |
|---|---|---|
| *West North Central-South Atlantic* | 0.392 | 1.27 |
| *South Atlantic-Middle Atlantic* | 0.351 | 1.36 |
| *Mountain West-East North Central* | 0.268 | 1.38 |
| *East South Central-West South Central* | 0.134 | 3.1 |
| *Middle Atlantic-Mountain West* | 0.117 | 2.13 |
| *West North Central-Middle Atlantic* | 0.111 | 2.62 |
| *Middle Atlantic-East North Central* | 0.0221 | 3.51 |
| *East North Central-New England* | 0.0203 | 3.15 |
| *South Atlantic-Mountain West* | 0.00271 | 3.49 |
| *West North Central-Mountain West* | <0.001 | 4.75 |
| *South Atlantic-East North Central* | <0.001 | 4.86 |
| *Mountain West-New England* | <0.001 | 4.53 |
| *West North Central-East North Central* | <0.001 | 6.13 |
| *Middle Atlantic-New England* | <0.001 | 6.66 |
| *West North Central-New England* | <0.001 | 9.28 |
| *South Atlantic-New England* | <0.001 | 8.02 |
| *West South Central-West North Central* | <0.001 | 11.59 |
| *East South Central-West North Central* | <0.001 | 14.7 |
| *West South Central-South Atlantic* | <0.001 | 12.86 |
| *West South Central-Middle Atlantic* | <0.001 | 14.21 |
| *East South Central-South Atlantic* | <0.001 | 15.96 |
| *East South Central-Middle Atlantic* | <0.001 | 17.32 |
| *East South Central-Mountain West* | <0.001 | 19.45 |
| *East South Central-East North Central* | <0.001 | 20.83 |
| *West South Central-Mountain West* | <0.001 | 16.35 |
| *West South Central-East North Central* | <0.001 | 17.72 |
| *New England-Pacific West* | <0.001 | 12.01 |
| *East South Central-New England* | <0.001 | 23.98 |
| *East North Central-Pacific West* | <0.001 | 15.16 |
| *West South Central-New England* | <0.001 | 20.88 |
| *Middle Atlantic-Pacific West* | <0.001 | 18.67 |
| *West North Central-Pacific West* | <0.001 | 21.29 |
| *East South Central-Pacific West* | <0.001 | 35.99 |
| *South Atlantic-Pacific West* | <0.001 | 20.02 |
| *Mountain West-Pacific West* | <0.001 | 16.54 |
| *West South Central-Pacific West* | <0.001 | 32.88 |

**Supplementary Table 5:** Membership and visit proportions by age and region.

| AGE GROUP | REGION | AVERAGE VISIT PROPORTION (95% CI) | AVERAGE MEMBERSHIP PROPORTION (95% CI) |
|---|---|---|---|
| **0-4** | Midwest | 0.2 (0.2,0.2) | 0.06 (0.06,0.06) |
| **0-4** | Northeast | 0.2 (0.2,0.2) | 0.05 (0.05,0.05) |
| **0-4** | South | 0.26 (0.25,0.27) | 0.05 (0.05,0.05) |
| **0-4** | West | 0.18 (0.17,0.18) | 0.06 (0.06,0.06) |
| **5-9** | Midwest | 0.33 (0.32,0.34) | 0.07 (0.07,0.07) |
| **5-9** | Northeast | 0.35 (0.35,0.36) | 0.06 (0.06,0.07) |
| **5-9** | South | 0.32 (0.31,0.33) | 0.06 (0.06,0.07) |
| **5-9** | West | 0.28 (0.27,0.28) | 0.07 (0.07,0.07) |
| **10-19** | Midwest | 0.21 (0.2,0.21) | 0.16 (0.16,0.16) |
| **10-19** | Northeast | 0.22 (0.21,0.22) | 0.15 (0.15,0.15) |
| **10-19** | South | 0.19 (0.18,0.19) | 0.15 (0.15,0.15) |
| **10-19** | West | 0.22 (0.22,0.22) | 0.16 (0.15,0.16) |
| **20-29** | Midwest | 0.09 (0.08,0.09) | 0.13 (0.13,0.14) |
| **20-29** | Northeast | 0.08 (0.07,0.08) | 0.13 (0.12,0.14) |
| **20-29** | South | 0.08 (0.08,0.08) | 0.13 (0.13,0.14) |
| **20-29** | West | 0.12 (0.12,0.13) | 0.14 (0.13,0.14) |
| **30-39** | Midwest | 0.09 (0.09,0.1) | 0.14 (0.14,0.14) |
| **30-39** | Northeast | 0.07 (0.07,0.08) | 0.14 (0.14,0.14) |
| **30-39** | South | 0.08 (0.07,0.08) | 0.15 (0.15,0.15) |
| **30-39** | West | 0.11 (0.11,0.11) | 0.15 (0.15,0.15) |
| **40-49** | Midwest | 0.04 (0.04,0.05) | 0.17 (0.16,0.17) |
| **40-49** | Northeast | 0.04 (0.04,0.05) | 0.18 (0.17,0.19) |
| **40-49** | South | 0.04 (0.04,0.04) | 0.18 (0.18,0.18) |
| **40-49** | West | 0.05 (0.05,0.06) | 0.17 (0.17,0.17) |
| **50-59** | Midwest | 0.02 (0.02,0.02) | 0.2 (0.19,0.2) |
| **50-59** | Northeast | 0.02 (0.02,0.02) | 0.2 (0.2,0.21) |
| **50-59** | South | 0.02 (0.02,0.03) | 0.19 (0.19,0.2) |
| **50-59** | West | 0.03 (0.03,0.03) | 0.19 (0.18,0.19) |
| **60-69** | Midwest | 0.01 (0.01,0.01) | 0.07 (0.07,0.08) |
| **60-69** | Northeast | 0.01 (0.01,0.01) | 0.08 (0.07,0.08) |
| **60-69** | South | 0.01 (0.01,0.01) | 0.07 (0.07,0.07) |
| **60-69** | West | 0.01 (0.01,0.02) | 0.07 (0.06,0.07) |

**Supplementary Table 6: Region Difference Comparisons by Quarter**

Quarter 1 = January – March, Quarter 2 = April – June, Quarter 3 = July – September, Quarter 4 = October - December

| Comparison | Quarter | Absolute Difference in Visits per 1000 People (95% CI) |
|---|---|---|
| **Northeast-Midwest** | 1 | 0.14 (-0.51-0.79) |
| **Northeast-Midwest** | 2 | 0.93 (0.61-1.26) |
| **Northeast-Midwest** | 3 | 0.41 (0.23-0.59) |
| **Northeast-Midwest** | 4 | 0.51 (0.25-0.78) |
| **South-Midwest** | 1 | 1.97 (1.41-2.54) |
| **South-Midwest** | 2 | 2.2 (1.8-2.6) |
| **South-Midwest** | 3 | 2.1 (1.92-2.29) |
| **South-Midwest** | 4 | 3.38 (2.97-3.8) |
| **South-Northeast** | 1 | 2.11 (1.58-2.65) |
| **South-Northeast** | 2 | 1.27 (0.93-1.61) |
| **South-Northeast** | 3 | 2.51 (2.25-2.78) |
| **South-Northeast** | 4 | 3.9 (3.55-4.25) |
| **West-Midwest** | 1 | 5.02 (3.98-6.07) |
| **West-Midwest** | 2 | 2.93 (2.49-3.37) |
| **West-Midwest** | 3 | 1.34 (1.09-1.59) |
| **West-Midwest** | 4 | 2.53 (2.27-2.79) |
| **West-Northeast** | 1 | 4.88 (4.13-5.64) |
| **West-Northeast** | 2 | 3.87 (3.36-4.37) |
| **West-Northeast** | 3 | 0.93 (0.76-1.1) |
| **West-Northeast** | 4 | 2.02 (1.7-2.33) |
| **West-South** | 1 | 7 (5.98-8.01) |
| **West-South** | 2 | 5.14 (4.59-5.68) |
| **West-South** | 3 | 3.44 (3.06-3.83) |
| **West-South** | 4 | 5.91 (5.36-6.46) |

**Supplementary Table 7:** State sinusoid phases with confidence intervals

| *State* | *Phase (95% CI)* |
|---|---|
| *Alabama* | 1.4 (1.09-1.7) |
| *Arizona* | 1.82 (1.49-2.15) |
| *Arkansas* | 1.37 (0.99-1.74) |
| *California* | 2.35 (2.06-2.63) |
| *Colorado* | 1.63 (1.39-1.88) |
| *Connecticut* | 2.61 (2.42-2.8) |
| *Delaware* | 2.41 (2.1-2.73) |
| *Florida* | 2.17 (1.78-2.56) |
| *Georgia* | 1.55 (1.2-1.91) |
| *Idaho* | 1.45 (1.17-1.73) |
| *Illinois* | 2.22 (1.97-2.47) |
| *Indiana* | 1.97 (1.74-2.21) |
| *Iowa* | 2.04 (1.78-2.3) |
| *Kansas* | 1.9 (1.6-2.2) |
| *Kentucky* | 1.65 (1.34-1.96) |
| *Louisiana* | 0.81 (0.41-1.21) |
| *Maine* | 2.38 (2.12-2.65) |
| *Maryland* | 2.5 (2.26-2.73) |
| *Massachusetts* | 2.7 (2.5-2.89) |
| *Michigan* | 2.42 (2.15-2.69) |
| *Minnesota* | 2.38 (2.12-2.64) |
| *Mississippi* | 0.86 (0.43-1.3) |
| *Missouri* | 1.79 (1.56-2.01) |
| *Montana* | 1.45 (1.21-1.7) |
| *Nebraska* | 1.95 (1.58-2.31) |
| *Nevada* | 1.97 (1.43-2.52) |
| *New Hampshire* | 2.54 (2.33-2.75) |
| *New Jersey* | 2.66 (2.44-2.88) |
| *New Mexico* | 1.65 (1.39-1.91) |
| *New York* | 2.53 (2.35-2.71) |
| *North Carolina* | 2.17 (1.91-2.44) |
| *North Dakota* | 1.98 (1.54-2.43) |
| *Ohio* | 2.2 (1.95-2.44) |
| *Oklahoma* | 1.44 (1.11-1.78) |
| *Oregon* | 2.45 (2.12-2.78) |
| *Pennsylvania* | 2.53 (2.32-2.74) |
| *Rhode Island* | 2.64 (2.37-2.91) |
| *South Carolina* | -- |
| *South Dakota* | 1.98 (1.64-2.33) |
| *Tennessee* | 1.67 (1.4-1.95) |
| *Texas* | 1.57 (1.29-1.85) |
| *Utah* | 1.69 (1.49-1.88) |
| *Vermont* | 2.17 (1.79-2.55) |
| *Virginia* | 2.46 (2.2-2.71) |
| *Washington* | 2.31 (1.97-2.65) |
| *Washington DC* | 2.74 (2.22-3.27) |
| *West Virginia* | 1.88 (1.54-2.22) |
| *Wisconsin* | 2.44 (2.19-2.69) |
| *Wyoming* | 1.42 (1.07-1.78) |

**Supplemental Animation: Visits per 1000 people over time**.

The animation shows the number of visits per 1000 people in each state in the U.S. across each month and year of the observation period. States are shaded according to the number of visits, with more visits resulting in darker red color.

https://github.com/mkline1/StrepPharyngitis_Public/blob/main/figures/finalgif.gif

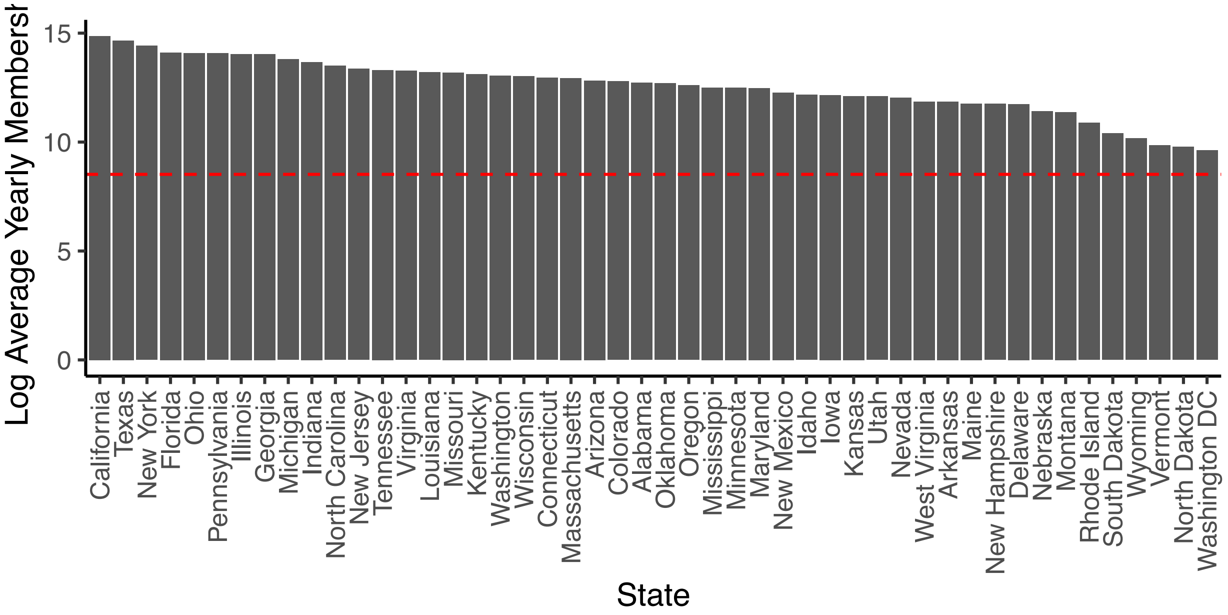


**Supplementary Figure 1**: Average membership in each state over the course of the study period. The dashed red line indicates the predetermined quality threshold of an average of 5,000 members per year.

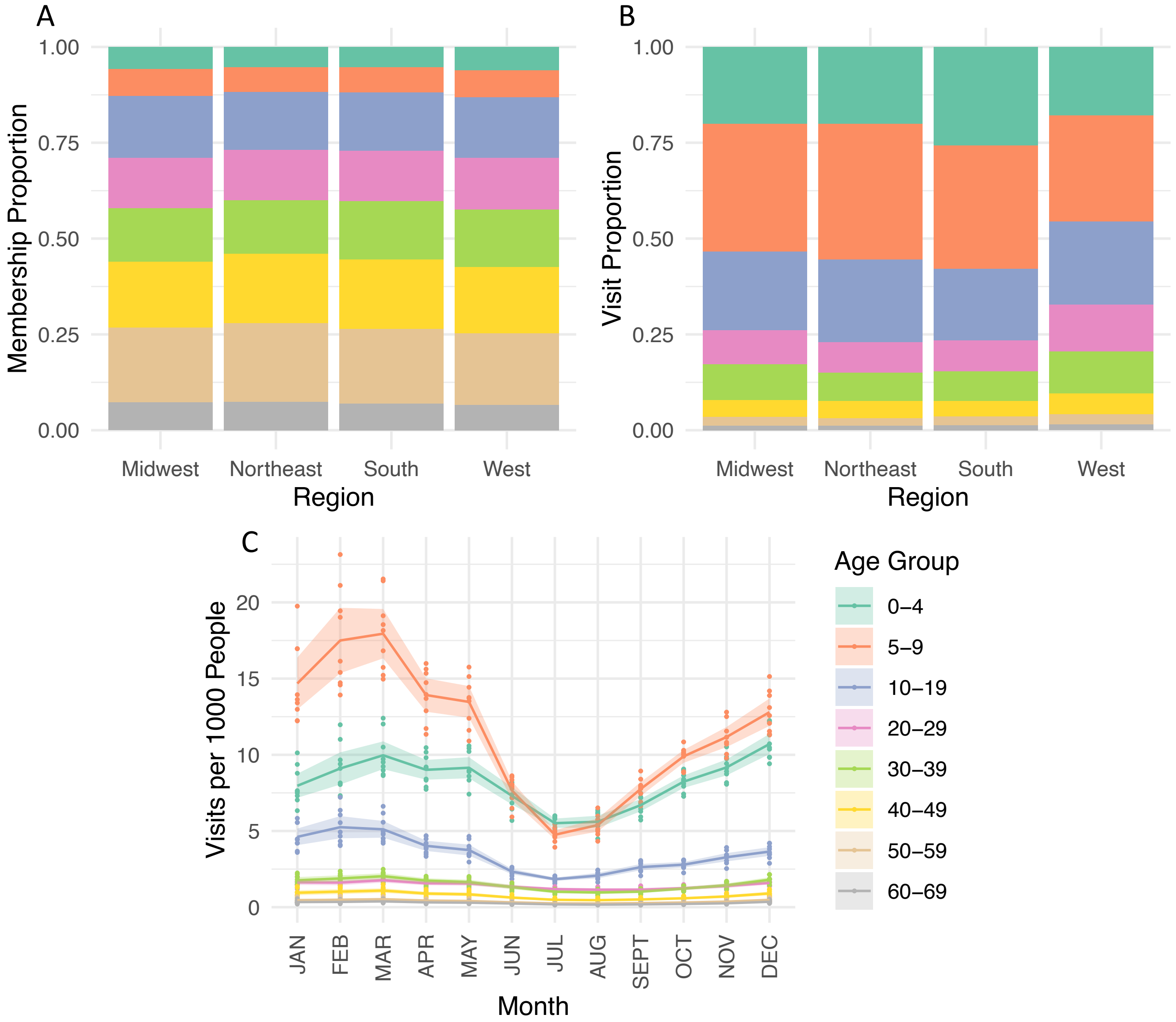


**Supplementary Figure 2:** Age distributions of the member and visit populations. Panel A: The proportion of members in each region that fall into each age group in an average year in the observation period. Panel B: The proportion of GAS pharyngitis visits in each region that fall into each age category. Panel C: The average number of visits per 1000 members in each age group in each month over the course of an average year. Shading represents 95% confidence intervals; points are individual year observations.

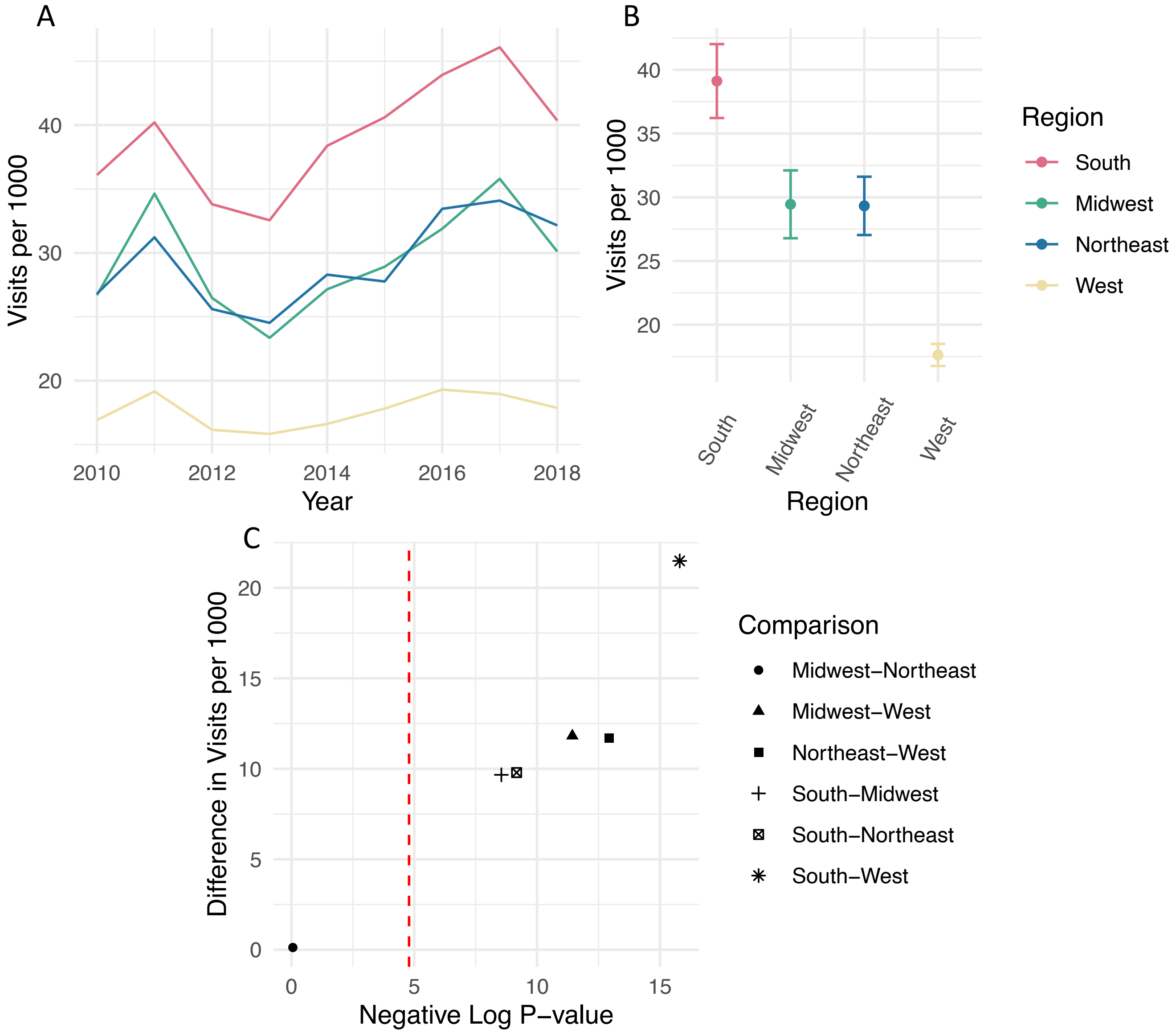


**Supplementary Figure 3:** Visits per 1000 people per year in each region. Panel A: Visits per 1000 people in each region over the 9-year observation period. Panel B: Average visits per 1000 people per year in each region with brackets showing 95% confidence intervals representing year-to-year variability. Panel C: Comparisons between each region pair. The y-axis shows the difference in average visits per 1000 people per year. The x-axis shows the negative log p-value from Welch's two-sample t-test comparing the 9 observations from each region. The dashed line indicates the significance threshold of 0.05 corrected for multiple hypothesis testing with the Bonferroni correction, where points to the right of the line are statistically significant and points to the left are not.

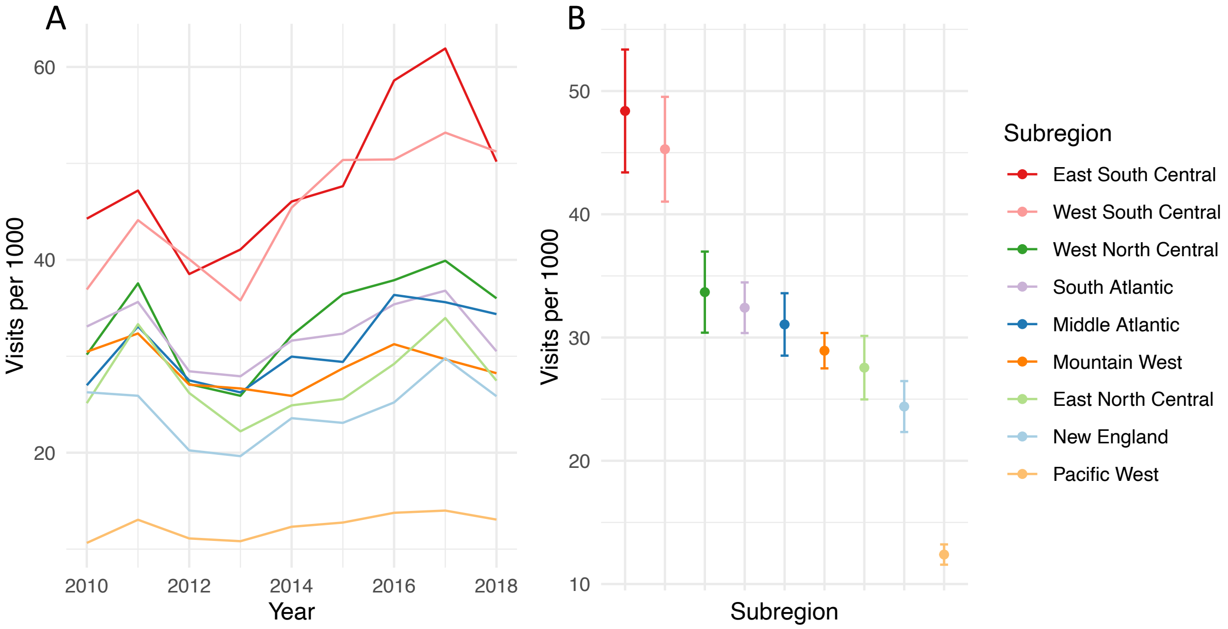


**Supplementary Figure 4:** Visits per 1000 people per year in each subregion. Panel A: Visits per 1000 people over the 9-year observation period. Panel B**:** Average visits per 1000 people per year in each subregion with brackets showing 95% confidence intervals representing year-to-year variability.

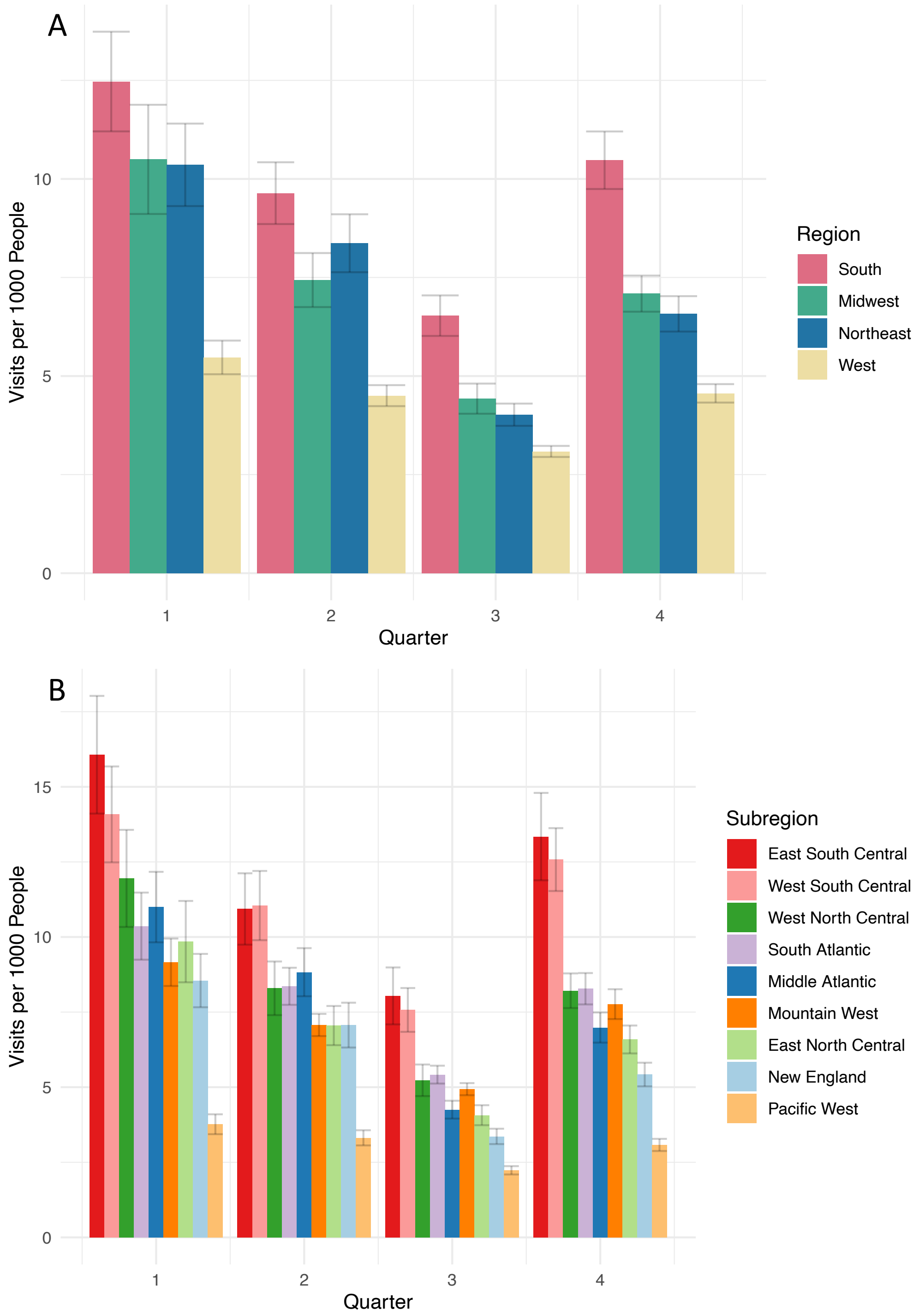


**Supplementary Figure 5:** Quarterly average visits per 1,000 people. Quarter 1 is January-March, Quarter 2 is April-June, Quarter 3 is July-September, Quarter 4 is October-December. Error bars are 95% confidence intervals assuming normally distributed errors. Panel A: By region. Panel B: By subregion.

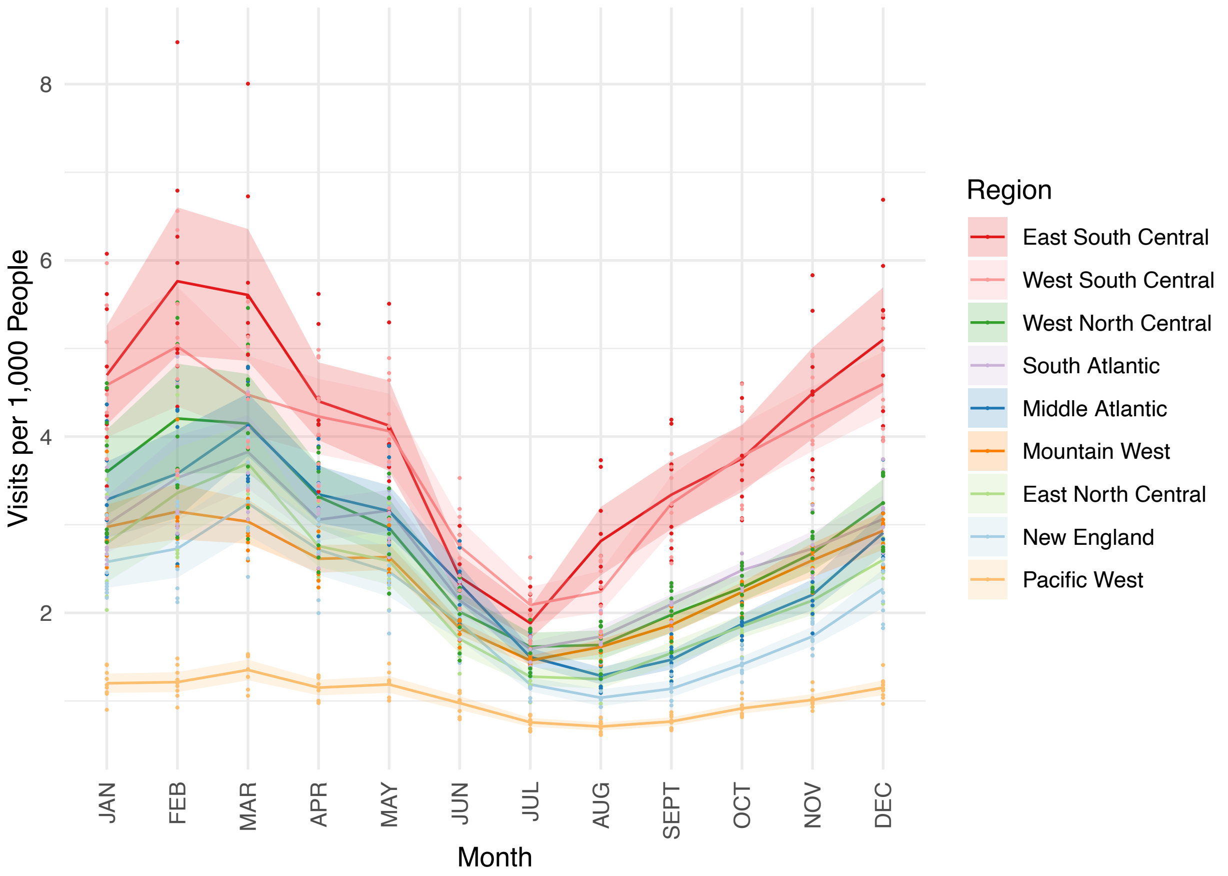


**Supplementary Figure 6: Average subregional GAS pharyngitis visit patterns over the course of the year**. The average number of visits per 1000 people in the database over the 9-year observation period for all age groups is plotted for each census subregion. Shading represents the 95% confidence intervals depicting year-to-year variation.

A

B

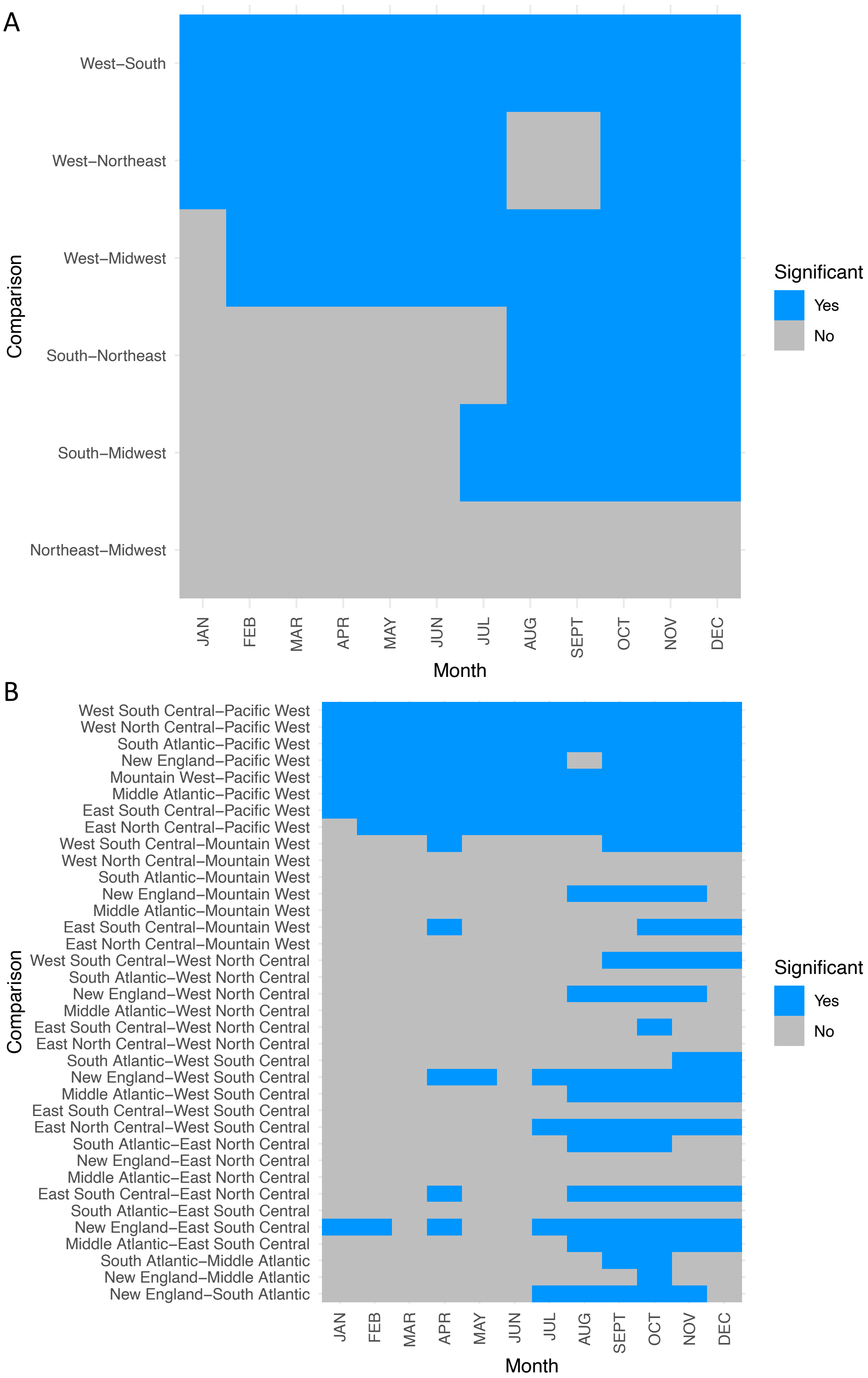
West–South
West–Northeast
West–Midwest
South–Northeast
South–Midwest
Northeast–Midwest
Comparison
JAN FEB MAR APR MAY JUN JUL AUG SEPT OCT NOV DEC
Month
Significant
Yes
No
West South Central–Pacific West
West North Central–Pacific West
South Atlantic–Pacific West
New England–Pacific West
Mountain West–Pacific West
Middle Atlantic–Pacific West
East South Central–Pacific West
East North Central–Pacific West
West South Central–Mountain West
West North Central–Mountain West
South Atlantic–Mountain West
New England–Mountain West
Middle Atlantic–Mountain West
East South Central–Mountain West
East North Central–Mountain West
West South Central–West North Central
South Atlantic–West North Central
New England–West North Central
Middle Atlantic–West North Central
East South Central–West North Central
East North Central–West North Central
South Atlantic–West South Central
New England–West South Central
Middle Atlantic–West South Central
East South Central–West South Central
East North Central–West South Central
South Atlantic–East North Central
New England–East North Central
Middle Atlantic–East North Central
East South Central–East North Central
South Atlantic–East South Central
New England–East South Central
Middle Atlantic–East South Central
South Atlantic–Middle Atlantic
New England–Middle Atlantic
New England–South Atlantic
Comparison
JAN FEB MAR APR MAY JUN JUL AUG SEPT OCT NOV DEC
Month
Significant
Yes
No

**Supplementary Figure 7: Monthly statistical comparison of Regional GAS Pharyngitis Visits.** Regions (Panel A) and subregions (Panel B) were compared via Welch's two sample t-test and significance was determined based on a significance level of 0.05 corrected using the Bonferroni correction for multiple hypothesis testing.

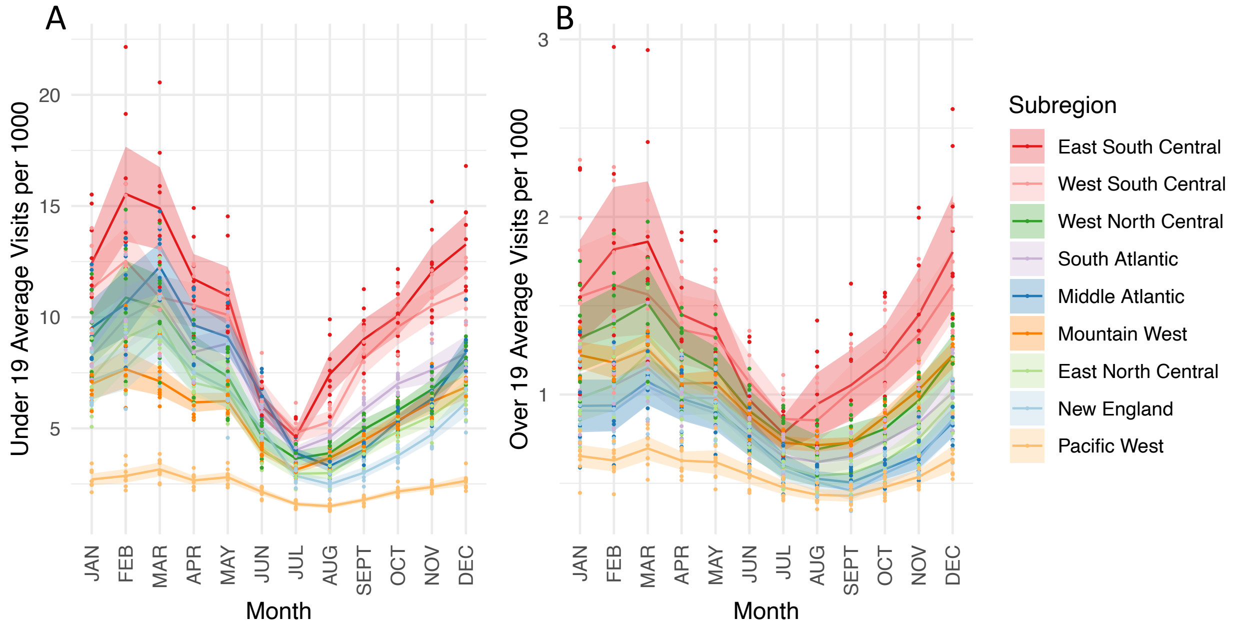


**Supplementary Figure 8: Monthly visits by subregion in the under 19 and over 19 age populations.** Panel A: average number of visits per 1000 under 19-year-old people in the database over the 9-year observation period for all age groups is plotted for each census subregion. Panel B: average number of visits per 1000 over 19-year-old people in the database over the 9-year observation period for all age groups is plotted for each census subregion (note the difference in the vertical scales). Shading represents the 95% confidence intervals depicting year-to-year variation.

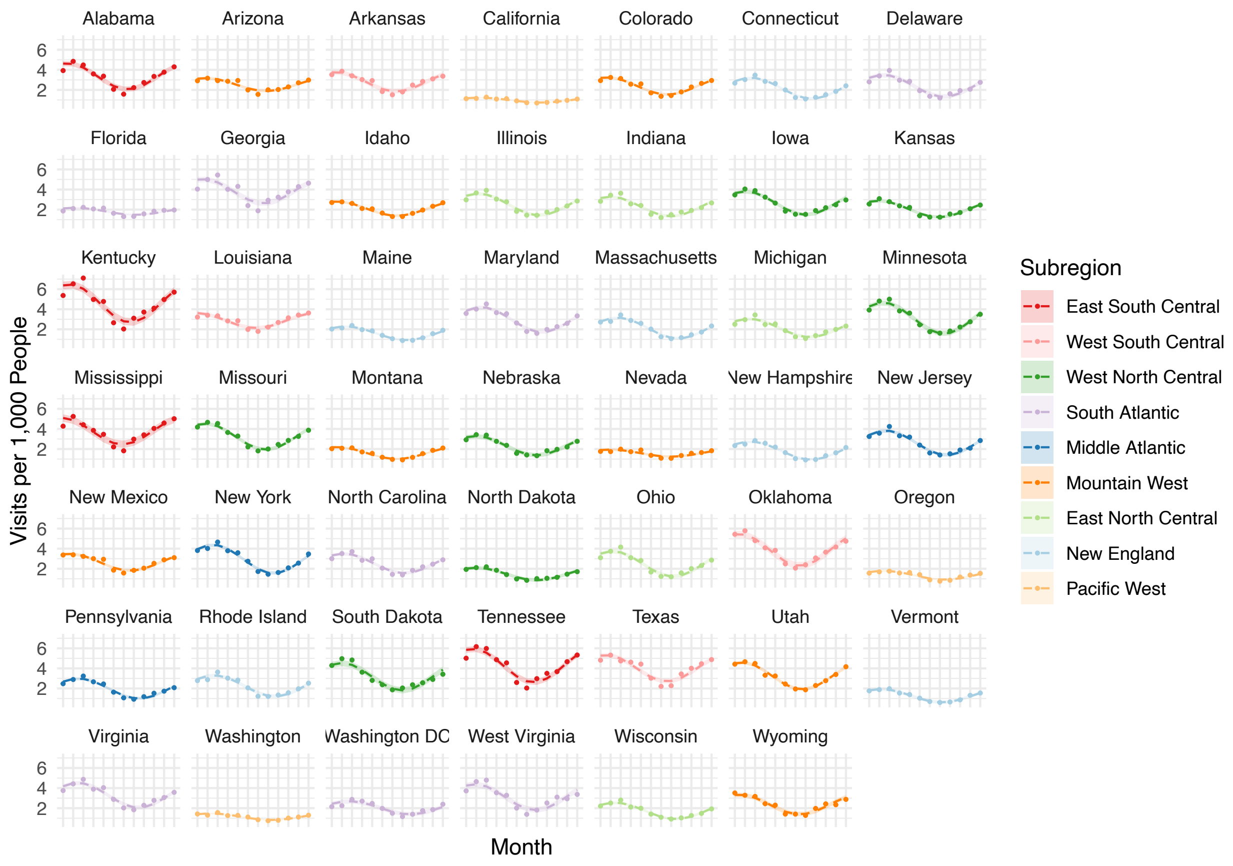


**Supplementary Figure 9: Individual state sinusoidal fits used to generate Figure 2**. Points are average GAS pharyngitis visits in that month. Dashed lines represent sinusoid fits, and shading represents 95% confidence intervals around sinusoid fits assuming normally distributed errors.

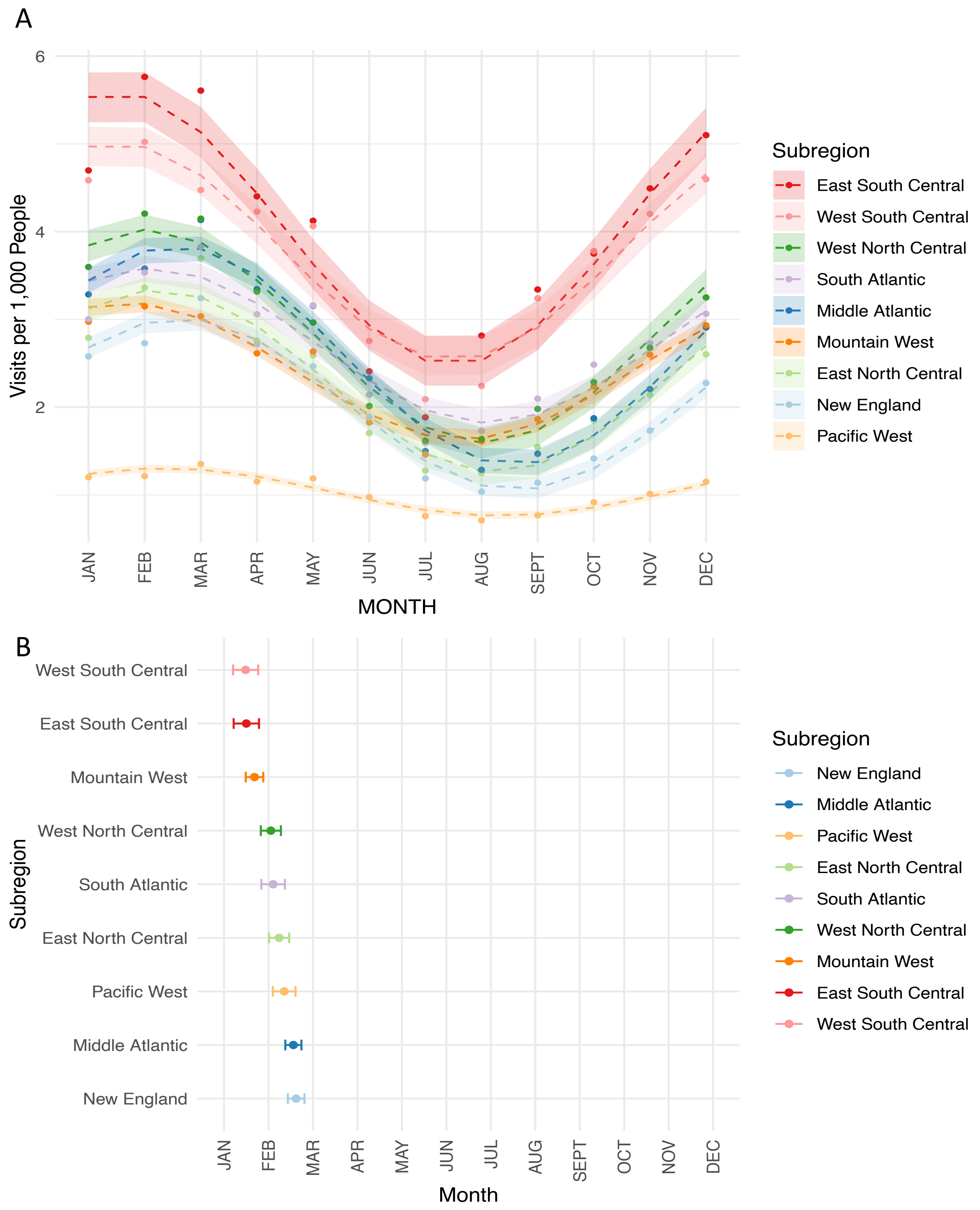


**Supplementary Figure 10**: **Subregion sinusoidal fits**. Panel A: GAS pharyngitis visit predictions according to subregional sinusoidal fitting. Points represent average visits. Shading represents 95% confidence intervals assuming normally distributed errors. Panel B: Sinusoidal fit phases in temporal order. Brackets represent 95% confidence intervals around the phase estimations.

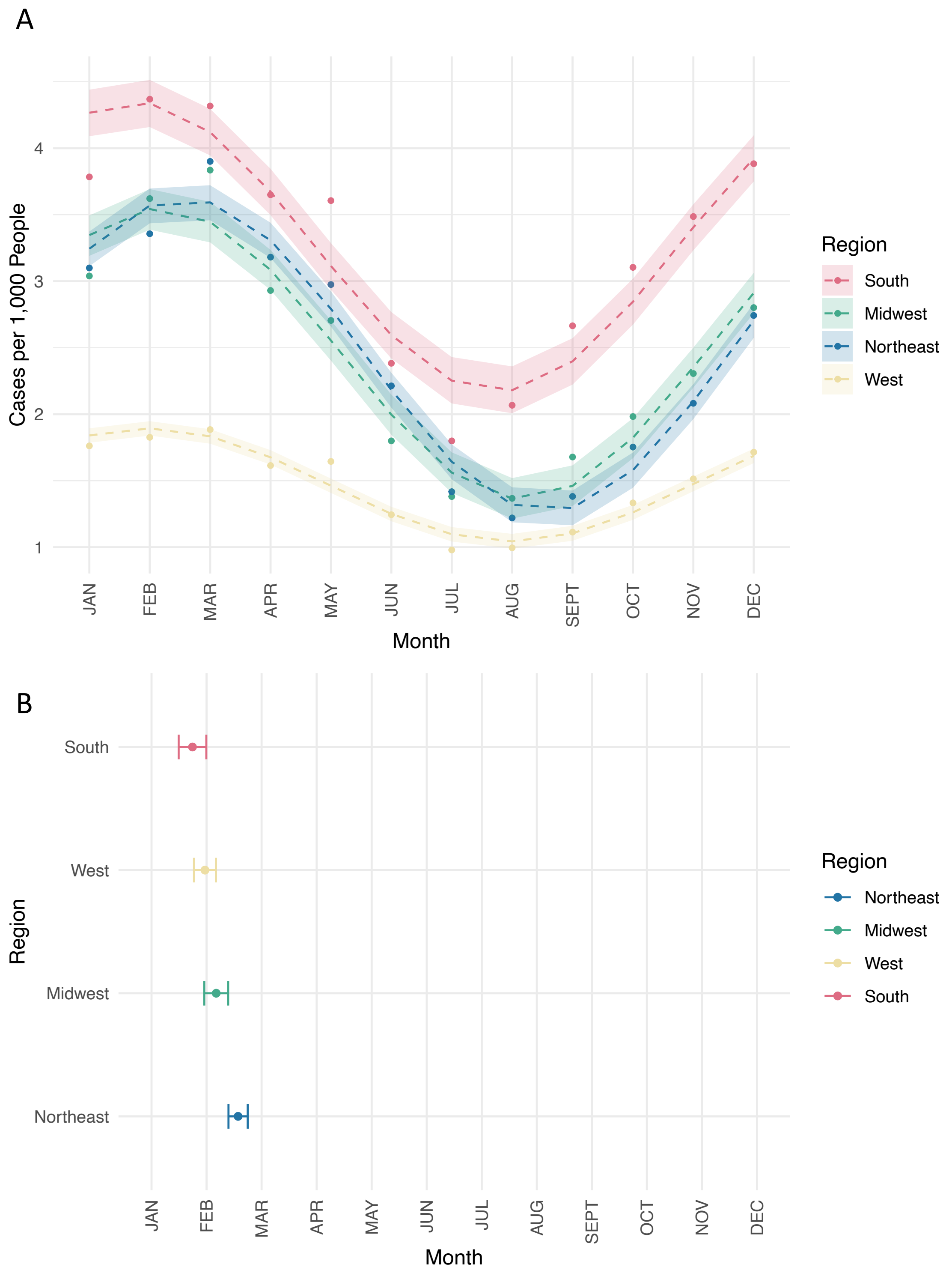


**Supplementary Figure 11: Region sinusoidal fits**. Panel A: GAS pharyngitis visit predictions according to regional sinusoidal fitting. Points represent average visits. Shading represents 95% confidence intervals assuming normally distributed errors. Panel B: Region sinusoidal fit phases in order. Brackets represent 95% confidence intervals around the phase estimations.

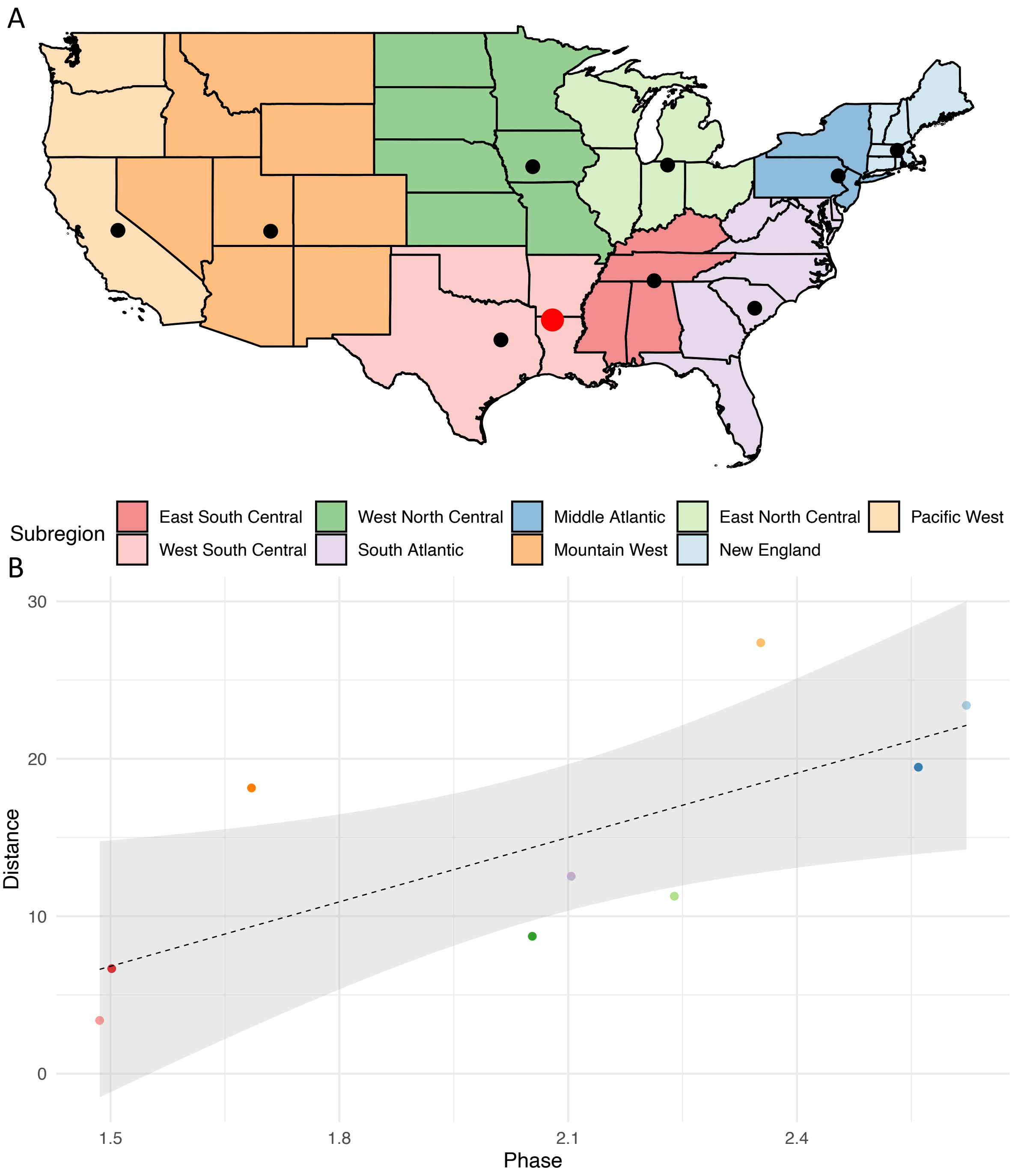


**Supplementary Figure 12: Correlation of subregion centroid with GAS Pharyngitis visit peak**. Panel A shows the population-weighted centroids of each subregion (black dots). The red point indicates the reference point for distance, which is a weighted average of the centroids of the East and West South Central subregions. Panel B shows sinusoid phase on the x-axis and distance from reference point on the y-axis. Points are colored according to their subregion. The dotted line is a linear regression and shading represents a 95% of the linear model's predictions. Pearson's correlation coefficient is $r = 0.723$.

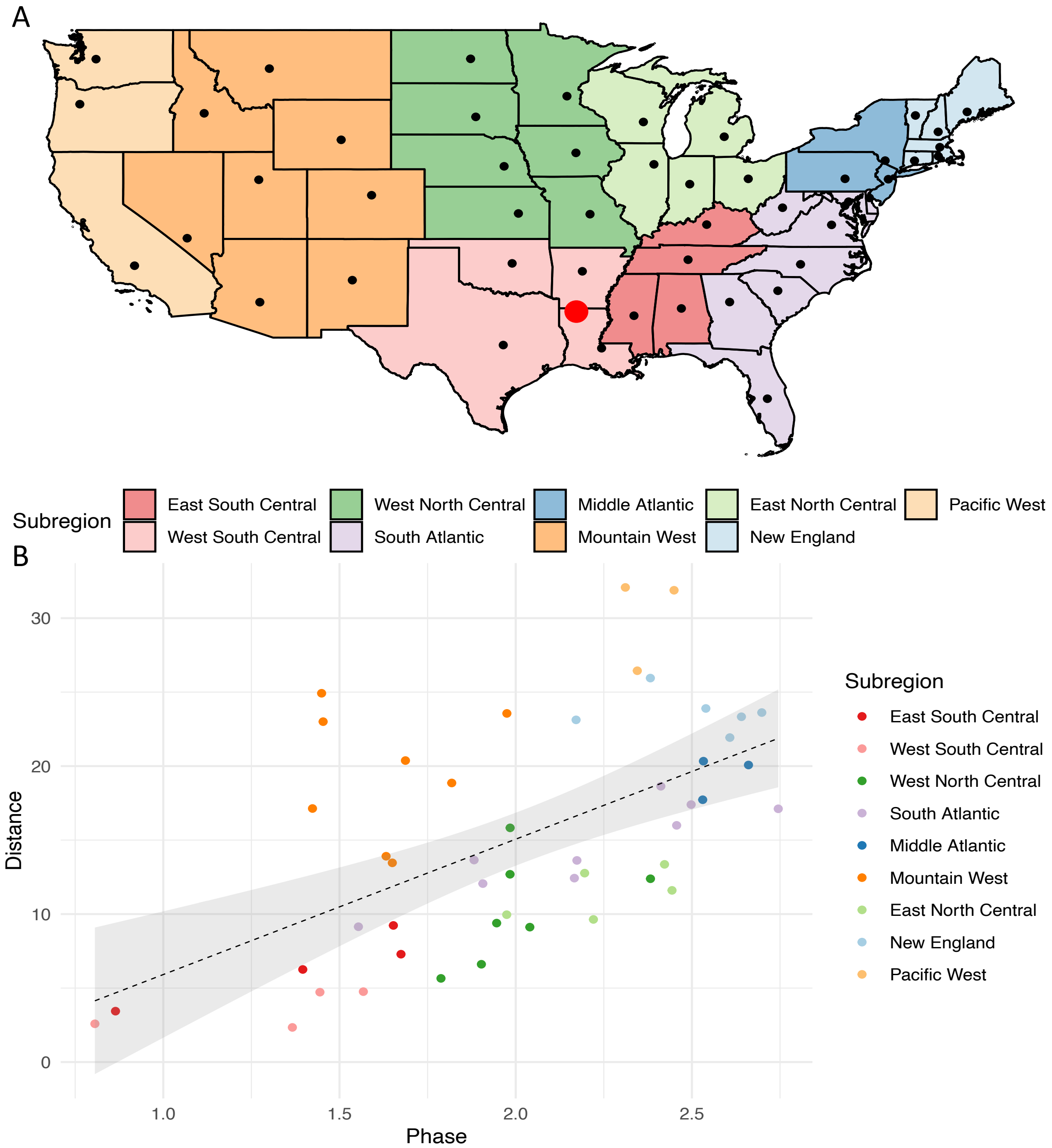


**Supplementary Figure 13**: **Correlation of state centroids with state GAS pharyngitis visit peak.** Panel A shows each state's centroid (black dots) according to the U.S. Census Bureau. The red point indicates the reference point for distance, which is a weighted average of the centroids of the East and West South Central subregions. Panel B shows state sinusoid phases plotted on the x-axis and distance from that state to the reference point on the y-axis. Points are colored according to their subregion. The dotted line is a linear regression and shading represents a 95% of the linear model's predictions. Pearson's correlation coefficient is $r = 0.575$.

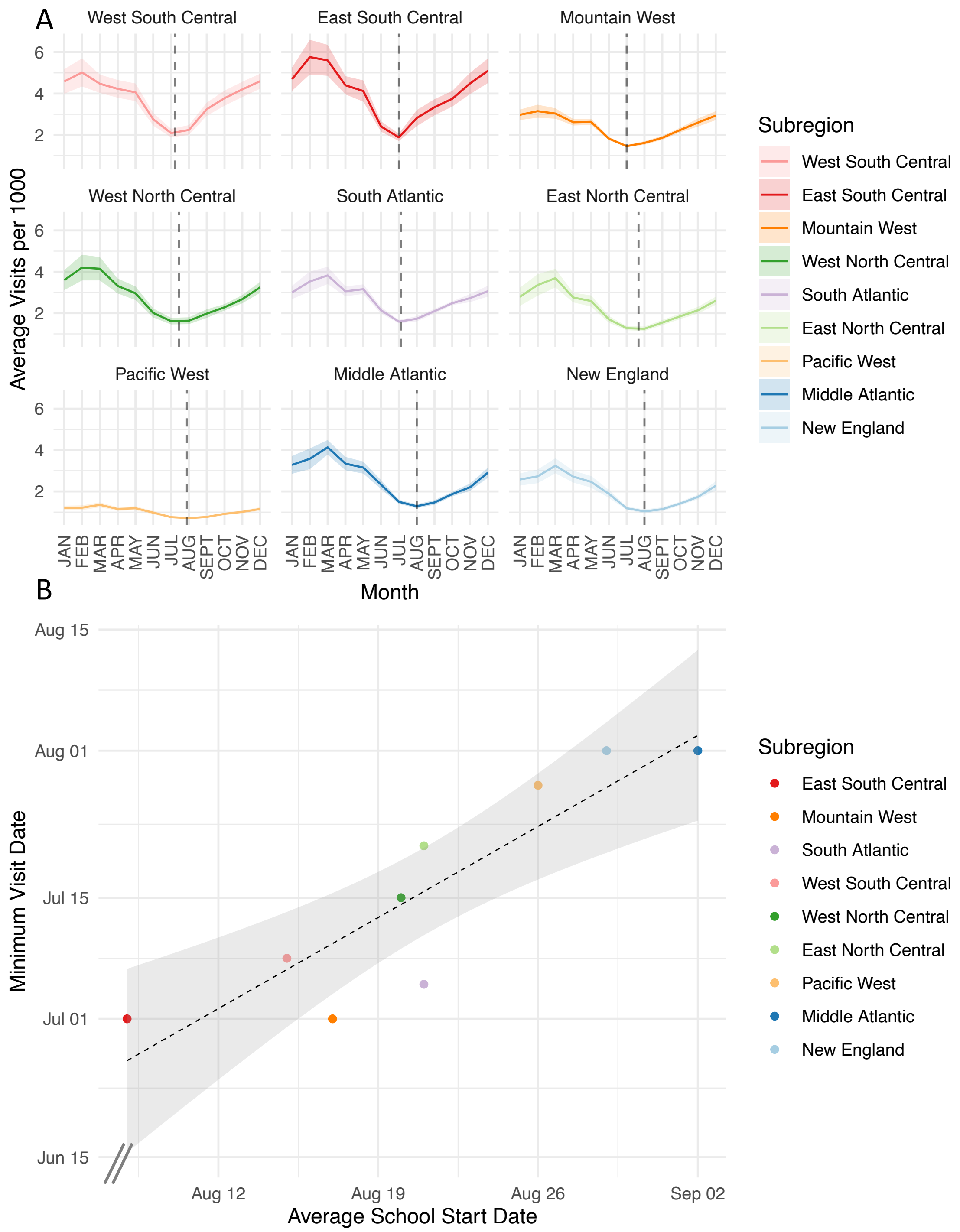


**Supplementary Figure 14**: **Comparison of subregion nadir visit date and school start date**. Panel A shows average minimum visit dates for each subregion visit trend with a dashed vertical line. Panel B shows average school start date plotted on the x-axis and minimum visit date is plotted on the y-axis. The dashed line represents a linear trend line with shading showing the 95% confidence interval of the linear model. Points are colored according to their subregion.

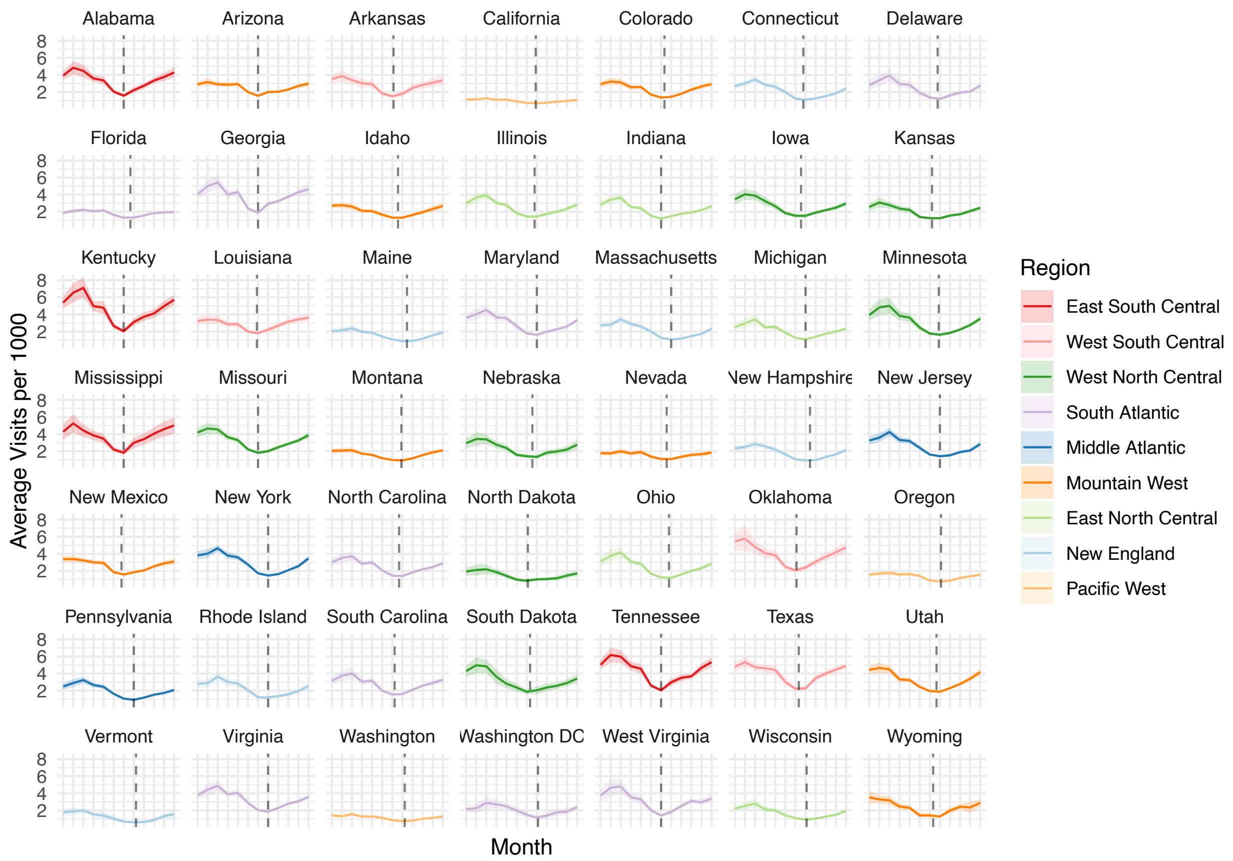


**Supplementary Figure 15: State minimum visit dates shown on top of state visit trends**. State trends averaged over the 9 years of observation are shown colored by that state's subregion. Shading represents 95% confidence intervals on the 9-year average. Dashed lines indicate the average minimum visit date for that state.

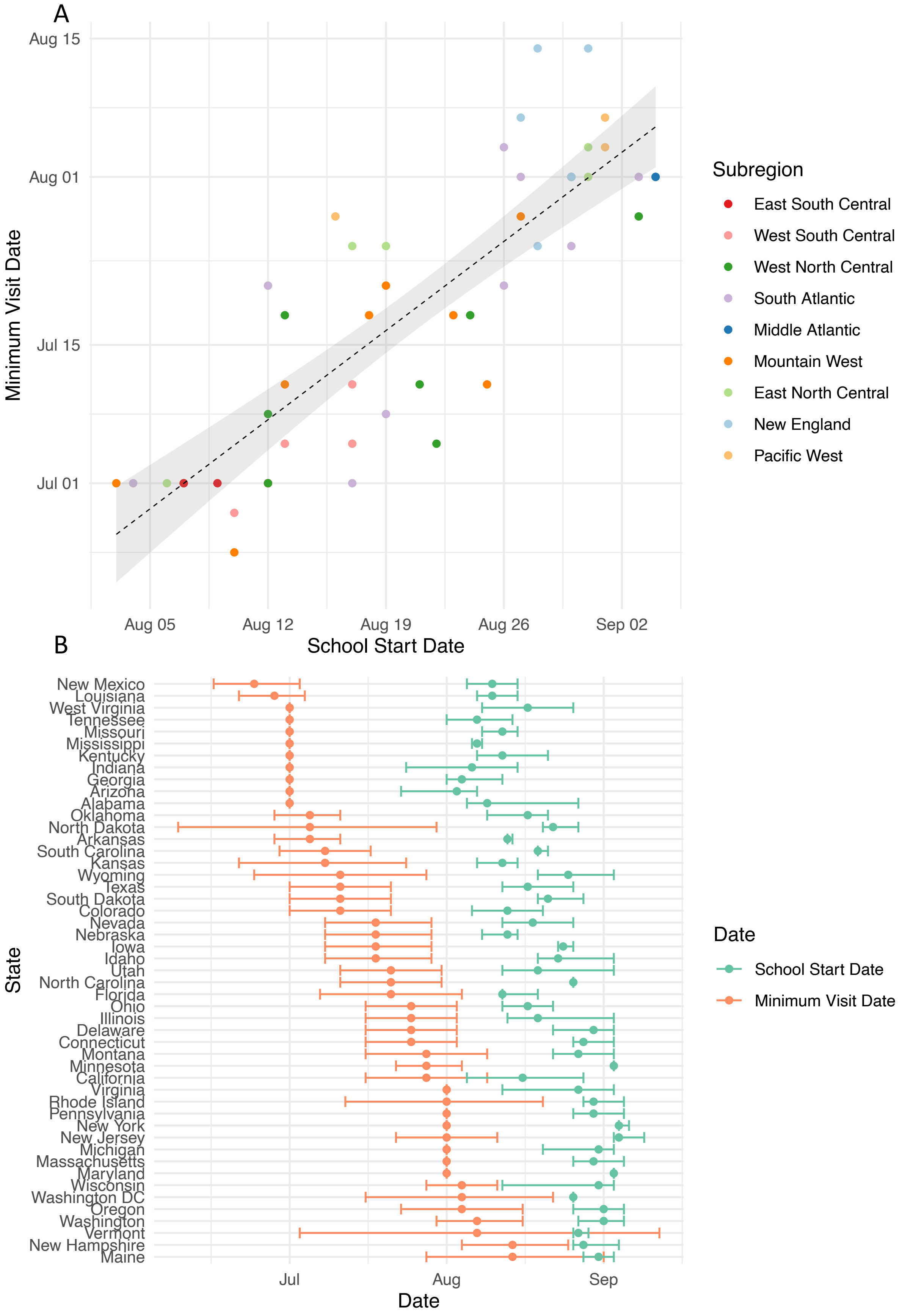
A
Aug 15
Aug 01
Jul 15
Jul 01
Minimum Visit Date
Aug 05
Aug 12
Aug 19
Aug 26
Sep 02
School Start Date
Subregion
East South Central
West South Central
West North Central
South Atlantic
Middle Atlantic
Mountain West
East North Central
New England
Pacific West
B
New Mexico
Louisiana
West Virginia
Tennessee
Missouri
Mississippi
Kentucky
Indiana
Georgia
Arizona
Alabama
Oklahoma
North Dakota
Arkansas
South Carolina
Kansas
Wyoming
Texas
South Dakota
Colorado
Nevada
Nebraska
Iowa
Idaho
Utah
North Carolina
Florida
Ohio
Illinois
Delaware
Connecticut
Montana
Minnesota
California
Virginia
Rhode Island
Pennsylvania
New York
New Jersey
Michigan
Massachusetts
Maryland
Wisconsin
Washington DC
Oregon
Washington
Vermont
New Hampshire
Maine
State
Jul
Aug
Sep
Date
Date
School Start Date
Minimum Visit Date

**Supplementary Figure 16: State minimum visit date compared with state school start date.** Panel A shows state-level correlation between average school start date in that state and average minimum visit date (r = 0.84, 95% CI: 0.82-0.86). States are colored by their corresponding subregion. Panel B shows the state-level average minimum visit date plotted alongside average school start date. Error bars on average minimum visit dates represent 95% confidence intervals of the average assuming normally distributed errors. Error bars on school start dates show the range of school start dates for that state.

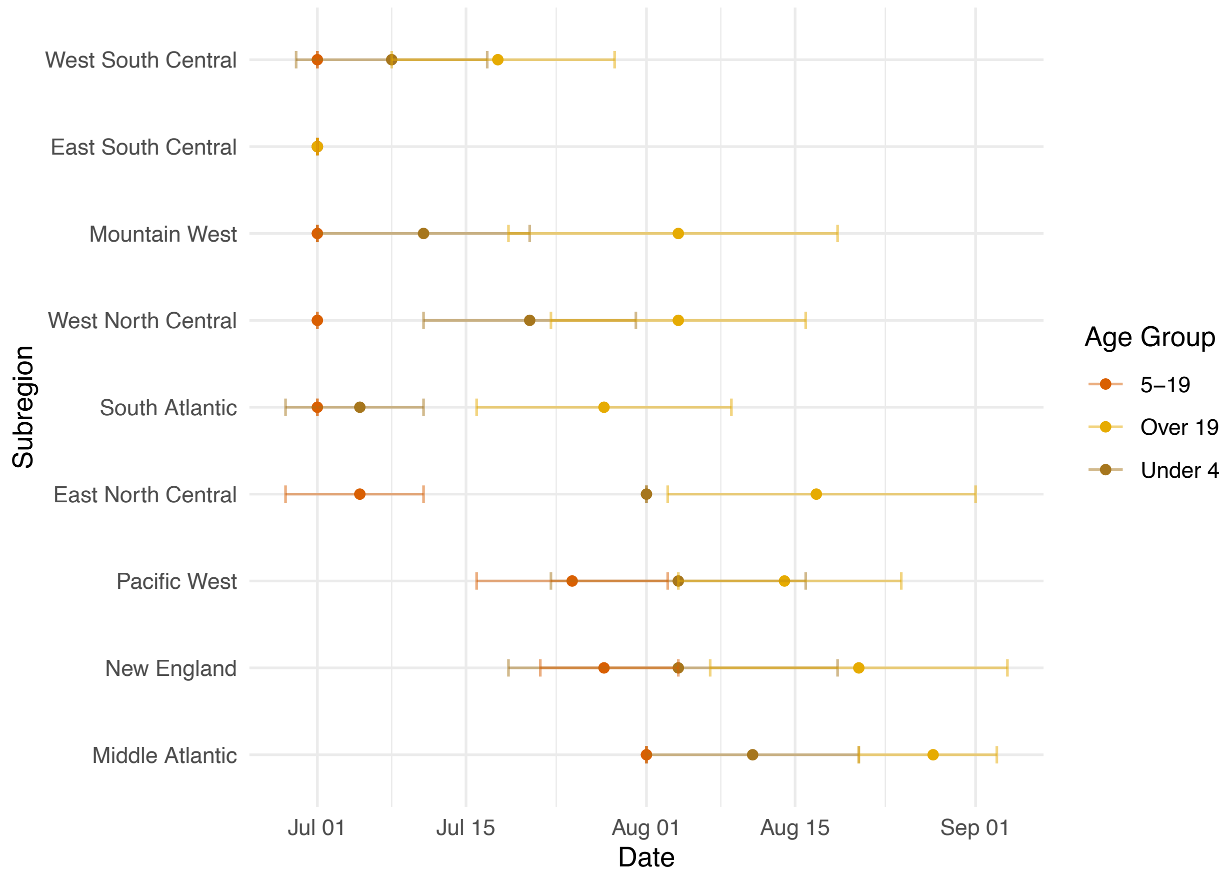


**Supplementary Figure 17:** Relationship between uptick dates between age groups. Average minimum visit dates with 95% confidence intervals assuming normally distributed errors in the 0-4, 5-19, and over 19 year old populations are shown alongside one another.
